# Longitudinal proteomic investigation of COVID-19 vaccination

**DOI:** 10.1101/2022.11.01.22281744

**Authors:** Yingrui Wang, Qianru Zhu, Rui Sun, Xiao Yi, Lingling Huang, Yifan Hu, Weigang Ge, Huanhuan Gao, Xinfu Ye, Yu Song, Li Shao, Yantao Li, Jie Li, Tiannan Guo, Junping Shi

**Author notes:** These authors contributed equally (Yingrui Wang, Qianru Zhu, Rui Sun, Xiao Yi).

## Abstract

Although the development of COVID-19 vaccines has been a remarkable success, the heterogeneous individual antibody generation and decline over time are unknown and still hard to predict. In this study, blood samples were collected from 163 participants who next received two doses of an inactivated COVID-19 vaccine (CoronaVac^®^) at a 28-day interval. Using TMT-based proteomics, we identified 1715 serum and 7342 peripheral blood mononuclear cells (PBMCs) proteins. We proposed two sets of potential biomarkers (seven from serum, five from PBMCs) using machine learning, and predicted the individual seropositivity 57 days after vaccination (AUC = 0.87). Based on the four PBMC’s potential biomarkers, we predicted the antibody persistence until 180 days after vaccination (AUC = 0.79). Our data highlighted characteristic hematological host responses, including altered lymphocyte migration regulation, neutrophil degranulation, and humoral immune response. This study proposed potential blood-derived protein biomarkers for predicting heterogeneous antibody generation and decline after COVID-19 vaccination, shedding light on immunization mechanisms and individual booster shot planning.

**Highlights:**

1. Longitudinal proteomics of PBMC and serum from individuals vaccinated with CoronaVac^®^.
2. Machine learning models predict neutralizing antibody generation and decline after COVID-19 vaccination.
3. The adaptive and the innate immune responses are stronger in the seropositive groups (especially in the early seropositive group).
4. Vaccine-induced immunity involves in lymphocyte migration regulation, neutrophil degranulation, and humoral immune response.

## Introduction

The global public health crisis and the social disruption caused by the coronavirus disease 2019 (COVID-19) pandemic have prompted the emergency use of speedily developed vaccines. As of October 2022, over 12 billion doses had been administered globally (WHO, 2022-10-25), although the vaccination distribution is significantly unbalanced (van der Graaf et al., 2022). Previous studies have reported that NAbs responses elicited by an inactivated vaccine (CoronaVac^®^) and an mRNA vaccine (BNT162b2) persisted for 6-8 months after full-schedule vaccination and declined to varying degrees (Falsey et al., 2021; Zeng et al., 2021). Therefore, multiple vaccine boosters and prolonged intervals between vaccine doses are needed to maintain the immunity against SARS-CoV-2 (Zhao et al., 2022), and could induce a robust humoral immune response (Ai et al., 2022). Several studies reported the dynamics of NAbs generation and the molecules dysregulation occurring after vaccinations (Liu et al., 2021; Wang et al., 2022). In a BMJ meta-analysis has shown that seroconversion rates and antibody titers after COVID-19 vaccines are significantly lower in immunocompromised patients than immunocompetent individuals (Lee et al., 2022), including immune-mediated inflammatory disorders, solid cancers, organ transplant recipients and hematological cancers. While to the best of our knowledge based on literature search, no study has systematically reported heterogeneous hematological host responses to vaccination in both PBMCs and serum. There is currently no known biomarker for predicting the effectiveness of vaccines.

In this study, we investigated the host response to Sinovac-CoronaVac^®^. Specifically, we analyzed the proteome of the peripheral blood mononuclear cells (PBMCs) and the sera of a vaccination recipients at different time points. We developed a method to predict the host responses to vaccination. Specifically, we predicted who cannot generate antibodies and whose NAbs tend to disappear earlier than six months after the vaccination. This information would help plan targeted boosters and decide the types and intervals of the vaccinations.

## Results

### Clinical and proteomics profiling before inactivated SARS-CoV-2 vaccination

Between January and February 2021, a total of 163 vaccination recipients were recruited in the discovery (N = 137) and the test (N = 26) cohorts (Figures 1A and 1B). The average age was 39.8 years in the discovery cohort and 41.6 years in the test cohort. Besides, most indexes of biochemical and hematology were not significantly different between the two cohorts. More details are shown in Tables 1-2, Table S1, and Figure S1. All the participants received the first dose of CoronaVac^®^ at day 0 (D0) and the second after 28 days (D28). The qualitative detection of SARS-CoV-2 NAbs and spike-specific IgG was done at D0, D28, day 57 (D57), and day 180 (D180). By D28, 19.6% of all participants (N = 32) were NAb seropositive (Group 2, the early seropositive group). By D57, the percentage of seropositive participants reached 88.3% (N = 144; Group 1+2, which included Group 1, the late seropositive group). The remaining 11.7% (N = 19) still had seronegative results (Group 0, the seronegative group). Within Group 1+2, 33.1% (N = 42) were still positive at D180 (Group 4, the persistently seropositive group), while the remaining ones became seronegative (Group 3) (Figure 1A). Besides, 10% of participants in Group 0 were IgG seropositive on day 28, which rising to 100% by day 57 and decreasing to 83% by day 180. However, 30% of participants in Group 1+2 were IgG seropositive on day 28, which rising to 100% by day 57 and decreasing to 88% by day 180 (Figure 1C). According to multivariable logistic regression analysis, we found that NAb titers at D28 were positively associated with seropositivity of neutralizing antibodies at D57 after adjusting age, sex, BMI and diastolic blood pressure. Then, we also identified that NAb titers at D28 and D57 could as independent predictors for seropositive of D180 after adjusting covariates (Figure S1). Blood samples were collected from all participants before their first vaccine dose, then at D28 and D57. Serum and PBMCs were extracted from all blood samples for proteomic profiling.

**Figure 1.**
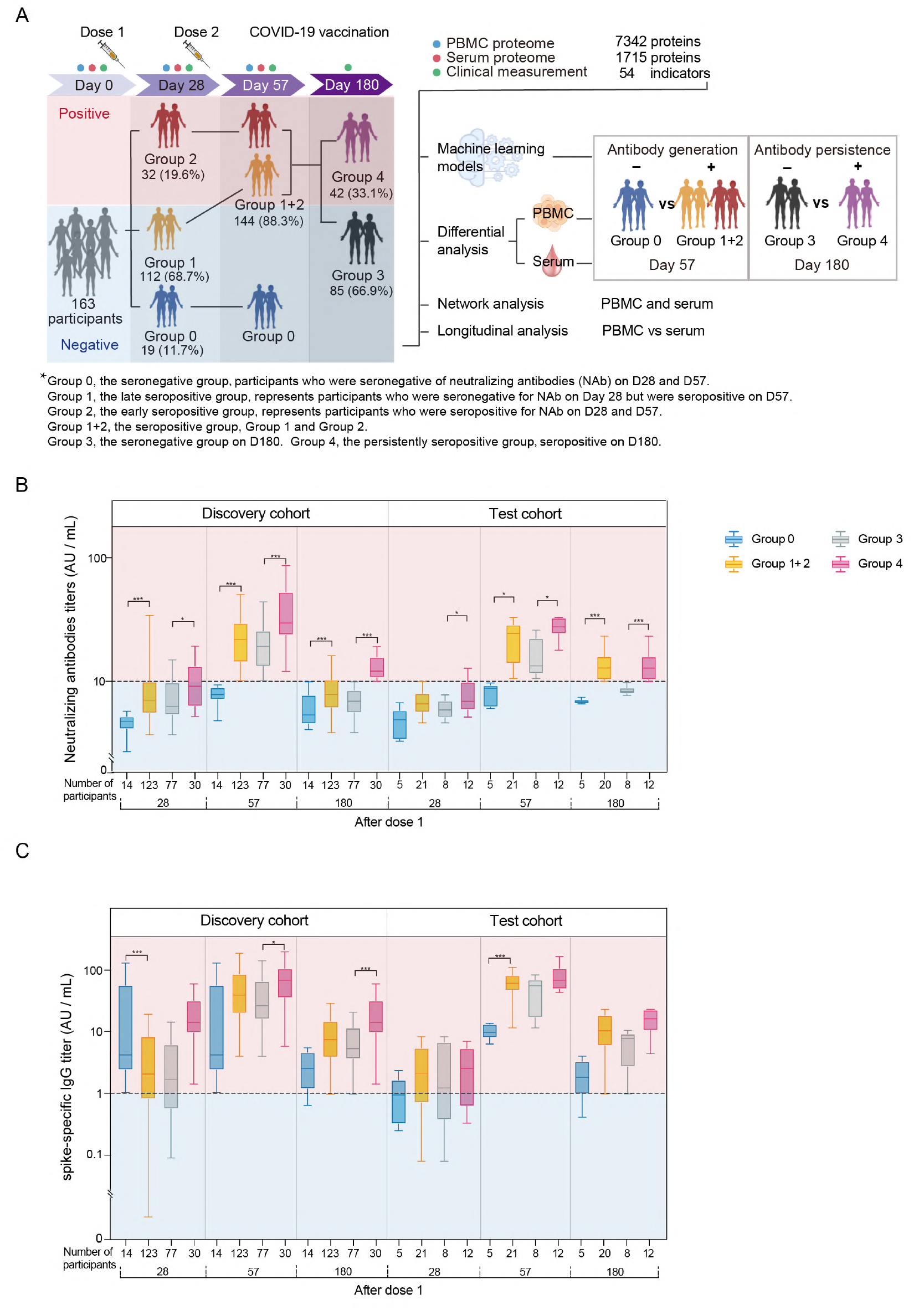
Study design and overview of clinical indicators. **(A)** Study design of the TMT labeling-based quantitative proteomics analysis of the PBMCs and sera samples. Vaccination recipients were vaccinated with two doses of 500 μl CoronaVac^®^, the first at D0 and the second at D28. Blood samples and PBMCs were collected at D0 (before vaccination), D28, and D57. Participants were divided into four groups based on the xenoreactivity of their NAbs on D28 and D57: Group 0, the seronegative group, included the participants that were seronegative on D28 and D57; Group 1, the late seropositive group, included the participants that were seronegative on Day 28 but seropositive on D57; Group 2, the early seropositive group, included the participants that were seropositive on D28 and D57. Groups 1 and 2 were combined into Group 1+2 to bring together all the seropositive participants. Group 1+2 was then divided into Group 3 (seronegative participants on D180) and Group 4 (seropositive participants on D180). **(B-C)** Antibody titers of neutralizing antibodies **(B)** and Spike-specific IgG **(C)** to live SARS-CoV-2 at different time points after vaccination. The horizontal line represents the threshold of specific response. The bars represent the median and IQR values of titers. Sample comparisons were tested by student t-test or Welch t-test. * Represents p < 0.05, ** represents p < 0.01, *** represents p < 0.001.

**Table 1.** Clinical metadata of the subjects for Group 0 and Group 1+2.

| Characteristics | Discovery cohort<br>(Cohort 1, N = 137) | Test cohort<br>(Cohort 2, N = 26) | p | Discovery cohort |  |  | Test cohort |  |  |
| --- | --- | --- | --- | --- | --- | --- | --- | --- | --- |
|  |  |  |  | Group 0<br>(N = 14) | Group 1+2 (N = 123) | p | Group 0<br>(N = 5) | Group 1+2<br>(N = 21) | p |
| Age (years) | 38.839 (11.806) | 41.615 (8.782) | 0.171 | 44.643<br>(12.258) | 38.179<br>(11.622) | 0.052 | 51.800 (5.495) | 39.191 (7.633) | 0.002 |
| Sex (male/female) | 43/94 | 16/10 | 0.141 | 6/8 | 37/86 | 0.329 | 5/0 | 11/10 | 0.123 |
| Body mass index (kg/m<sup>2</sup>) | 24.026 (3.770) | 25.314 (3.138) | 0.104 | 26.002 (4.465) | 23.801 (3.636) | 0.064 | 26.208 (4.955) | 25.097 (2.676) | 0.650 |
| Total bilirubin (μmol/L) | 19.883 (6.726) | 19.285 (5.663) | 0.671 | 19.650 (5.490) | 19.909 (6.871) | 0.892 | 20.080 (6.058) | 19.095 (5.706) | 0.734 |
| Albumin (g/L) | 48.457 (2.302) | 47.015 (2.706) | 0.005 | 47.979 (2.555) | 48.511 (2.277) | 0.414 | 45.820 (1.587) | 47.300 (2.865) | 0.281 |
| Alanine aminotransferase (U/L) | 20.479 (18.397) | 20.704<br>(10.932) | 0.952 | 17.607 (8.737) | 20.806<br>(19.186) | 0.540 | 19.840 (12.213) | 20.910 (10.923) | 0.849 |
| Aspartate aminotransferase (U/L) | 20.410 (9.150) | 22.054 (5.075) | 0.375 | 18.693 (5.496) | 20.606 (9.473) | 0.461 | 23.620 (6.460) | 21.681 (4.806) | 0.454 |
| Alkaline phosphatase (U/L) | 66.730 (20.253) | 69.462<br>(17.775) | 0.522 | 67.929<br>(19.828) | 66.594<br>(20.376) | 0.816 | 72.200 (12.696) | 68.810 (18.983) | 0.710 |
| γ-glutamyl transpeptidase (U/L) | 24.854 (21.930) | 22.692<br>(12.142) | 0.626 | 19.714<br>(10.542) | 25.439<br>(22.823) | 0.357 | 21.600 (7.956) | 22.952 (13.086) | 0.828 |
| LDL-cholesterol (mmol/L) | 3.118 (0.779) | 3.106 (0.883) | 0.941 | 2.796 (0.705) | 3.155 (0.782) | 0.102 | 3.264 (0.108) | 3.068 (0.982) | 0.665 |
| HDL-cholesterol (mmol/L) | 1.372 (0.326) | 1.220 (0.268) | 0.026 | 1.266 (0.182) | 1.384 (0.337) | 0.092 | 1.284 (0.163) | 1.205 (0.288) | 0.563 |
| Total cholesterol (mmol/L) | 4.965 (0.966) | 4.860 (0.983) | 0.611 | 4.557 (0.928) | 5.012 (0.963) | 0.103 | 4.984 (0.172) | 4.830 (1.094) | 0.760 |
| Triglyceride (mmol/L) | 1.017 (0.566) | 1.174 (0.645) | 0.207 | 1.089 (0.603) | 1.009 (0.564) | 0.618 | 0.962 (0.263) | 1.225 (0.701) | 0.424 |
| Glucose (mmol/L) | 4.146 (1.141) | 3.502 (0.826) | 0.007 | 4.210 (1.096) | 4.138 (1.150) | 0.825 | 3.398 (1.092) | 3.526 (0.782) | 0.762 |
| Creatinine (μmol/L) | 60.307 (29.087) | 67.923<br>(13.702) | 0.194 | 57.786<br>(17.375) | 60.594<br>(30.169) | 0.607 | 75.800 (13.046) | 66.048 (13.47) | 0.157 |
| Uric Acid (μmol/L) | 307.839 (95.341) | 343.385<br>(83.196) | 0.078 | 309.286<br>(52.889) | 307.675<br>(99.170) | 0.952 | 384.400<br>(75.075) | 333.619 (83.690) | 0.227 |
| CRP (mg/L) | 1.407 (3.782) | 1.206 (1.341) | 0.790 | 1.180 (1.460) | 1.442 (3.979) | 0.808 | 1.624 (1.984) | 1.235 (1.239) | 0.579 |
| Leukocytes (10<sup>9</sup>/L) | 6.203 (1.453) | 6.692 (1.517) | 0.120 | 2.164 (0.674) | 2.063 (0.591) | 0.594 | 6.320 (1.659) | 6.781 (1.511) | 0.552 |
| Platelets (10<sup>9</sup>/L) | 247.796 (57.838) | 271.500 (59.567) | 0.058 | 245.143 (62.954) | 248.098 (57.496) | 0.857 | 227.200 (53.742) | 282.048 (57.010) | 0.063 |
| Red blood cells (10<sup>9</sup>/L) | 6.203 (1.453) | 6.692 (1.517) | 0.120 | 4.742 (0.452) | 4.732 (0.630) | 0.953 | 5.320 (0.409) | 5.043 (0.605) | 0.344 |
| Lymphocytes (10<sup>9</sup>/L) | 2.074 (0.598) | 2.323 (0.513) | 0.048 | 2.164 (0.674) | 2.063 (0.591) | 0.552 | 2.060 (0.541) | 2.386 (0.499) | 0.209 |
| hemoglobin (g/L) | 140.985 (19.497) | 150.039 (20.166) | 0.032 | 138.143 (22.408) | 141.309 (19.215) | 0.567 | 160.800 (13.608) | 147.476 (20.868) | 0.190 |
| **Comorbidities, N (%)** |  |  |  |  |  |  |  |  |  |
| Hypertension | 13 (9) | 6 (23) | 0.005 | 1 (7) | 12 (10) | 0.092 | 2 (40) | 4 (19) | 0.558 |
| T2DM | 5 (4) | 0 (0) | >0.999 | 1 (7) | 4 (3) | 0.426 | 0 (0) | 0 (0) | >0.999 |
| MAFLD | 37 (27) | 13 (50) | 0.020 | 7 (50) | 30 (24) | 0.005 | 2 (40) | 11 (52) | >0.999 |
| **Seroconversion of neutralizing antibody to live SARS-CoV-2, N (%)** |  |  |  |  |  |  |  |  |  |
| Day 28 | 30 (22) | 2 (7) | 0.112 | 0 (0) | 30 (25) | 0.040 | 0 (0) | 2 (10) | >0.999 |
| Day 57 | 123 (90) | 21 (81) | 0.189 | 0 (0) | 123 (100) | >0.999 | 0 (0) | 21 (100) | <0.001 |
| Day 180 | 30 (25)<sup>a</sup> | 12 (48)<sup>b</sup> | 0.021 | 0 (0)<sup>c</sup> | 30 (28)<sup>d</sup> | >0.999 | 0 (0)<sup>e</sup> | 12 (60)<sup>f</sup> | 0.043 |
| **GMT of neutralizing antibody to live SARS-CoV-2 (AU/mL)** |  |  |  |  |  |  |  |  |  |
| Day 28 | 8.523 (5.368) | 6.789 (2.781) | 0.018 | 4.817 (1.185) | 8.945 (5.497) | <0.001 | 4.658 (1.379) | 7.296 (2.807) | .055 |
| Day 57 | 27.372 (28.155) | 23.309 (19.138) | 0.482 | 7.746 (1.293) | 29.606 (28.884) | <0.001 | 7.948 (1.64) | 26.967 (19.602) | .043 |
| Day 180 | 8.627 (4.315) | 10.795 (4.325) | 0.024 | 6.055 (1.976) | 8.94 (4.422) | <0.001 | 6.842 (0.343) | 11.784 (4.297) | <0.001 |

Data are shown as mean values (standard deviation within parentheses) and N (%). Pearson $\chi^2$ test or Fisher's exact test were used to analyze the categorical outcomes and student t-test or Welch t-test for continuous outcomes. Hypertension was defined as systolic blood pressure $\geq 140$ or diastolic blood pressure $\geq 90$ mmHg. T2DM: Type 2 diabetes mellitus, which was defined as fasting glucose $\geq 7.0$ mmol/L. MALFD: metabolic associated fatty liver disease. Superscripts a, b, c, d, e, and f: subjects left in each group were 120, 25, 13, 107, 5, and 20, respectively. Group 0, the seronegative group, included the participants that were seronegative on D28 and D57. Groups 1+2 were all the seropositive participants on D57.

**Table 2.** Clinical metadata of the subjects for Group 3 and Group 4.

| Characteristics | Discovery cohort<br>(Cohort 3, N = 107) | Test cohort<br>(Cohort 4, N = 20) | p | Discovery cohort |  | p | Test cohort |  | p |
| --- | --- | --- | --- | --- | --- | --- | --- | --- | --- |
|  |  |  |  | Group 3 (N = 77) | Group 4 (N = 30) |  | Group 3 (N = 8) | Group 4 (N = 12) |  |
| Age (years) | 40.019<br>(10.814) | 39.950 (6.970) | 0.971 | 40.130 (10.598) | 39.733 (11.531) | 0.866 | 40.625 (6.278) | 39.500 (7.634) | 0.734 |
| Sex (male/female) | 32/75 | 10/10 | 0.023 | 20/57 | 12/18 | 0.155 | 2/6 | 8/4 | 0.170 |
| Body mass index (kg/m<sup>2</sup>) | 23.965 (3.746) | 25.260 (2.638) | 0.142 | 23.879 (3.660) | 24.186 (4.014) | 0.705 | 26.738 (1.701) | 24.275 (2.745) | 0.037 |
| Total bilirubin (μmol/L) | 20.114 (7.080) | 18.735 (5.604) | 0.412 | 20.705 (6.615) | 18.597 (8.075) | 0.167 | 20.913 (6.399) | 17.283 (4.736) | 0.161 |
| Albumin (g/L) | 48.492 (2.245) | 47.175 (2.88) | 0.023 | 48.239 (2.191) | 49.140 (2.289) | 0.062 | 46.200 (2.183) | 47.825 (3.184) | 0.226 |
| Alanine aminotransferase (U/L) | 21.114<br>(19.713) | 21.310 (11.048) | 0.966 | 18.468 (16.533) | 27.907 (25.255) | 0.066 | 19.350 (12.294) | 22.617 (10.487) | 0.532 |
| Aspartate aminotransferase (U/L) | 20.425 (9.418) | 21.555 (4.895) | 0.602 | 19.136 (7.126) | 23.733 (13.243) | 0.08 | 20.738 (4.525) | 22.100 (5.248) | 0.556 |
| Alkaline phosphatase (U/L) | 66.720<br>(21.280) | 69.000 (19.456) | 0.657 | 66.065 (22.940) | 68.400 (16.492) | 0.612 | 59.875 (15.524) | 75.083 (19.988) | 0.087 |
| γ-glutamyl transpeptidase (U/L) | 26.178<br>(23.377) | 23.3 (13.326) | 0.595 | 24.208 (19.513) | 31.233 (31.030) | 0.164 | 18.375 (8.123) | 26.583 (15.341) | 0.184 |
| LDL-cholesterol (mmol/L) | 3.160 (0.800) | 3.076 (1.007) | 0.680 | 3.097 (0.769) | 3.320 (0.867) | 0.196 | 2.605 (0.655) | 3.389 (1.100) | 0.088 |
| HDL-cholesterol (mmol/L) | 1.370 (0.337) | 1.179 (0.27) | 0.018 | 1.406 (0.353) | 1.278 (0.276) | 0.076 | 1.188 (0.332) | 1.173 (0.235) | 0.912 |
| Total cholesterol (mmol/L) | 4.996 (0.935) | 4.821 (1.122) | 0.458 | 4.907 (0.910) | 5.223 (0.973) | 0.116 | 4.316 (0.698) | 5.157 (1.247) | 0.102 |
| Triglyceride (mmol/L) | 1.025 (0.596) | 1.245 (0.713) | 0.145 | 0.888 (0.451) | 1.375 (0.768) | 0.002 | 1.154 (0.657) | 1.305 (0.771) | 0.655 |
| Glucose (mmol/L) | 4.181 (1.222) | 3.593 (0.738) | 0.040 | 4.229 (1.275) | 4.060 (1.082) | 0.524 | 3.405 (0.708) | 3.718 (0.762) | 0.367 |
| Creatinine (μmol/L) | 60.009<br>(31.812) | 65.55 (13.621) | 0.446 | 59.610 (37.004) | 61.033 (10.440) | 0.836 | 63.000 (10.637) | 67.250 (15.51) | 0.509 |
| Uric Acid (μmol/L) | 301.122<br>(100.483) | 331.650<br>(85.363) | 0.205 | 288.299<br>(93.366) | 334.033<br>(111.819) | 0.034 | 349.000<br>(91.377) | 320.083 (83.115) | 0.473 |
| CRP (mg/L) | 1.224 (2.529) | 1.136 (1.209) | 0.879 | 1.270 (2.904) | 1.106 (1.151) | 0.764 | 1.084 (0.997) | 1.170 (1.374) | 0.881 |
| Leukocytes (10<sup>9</sup>/L) | 6.173 (1.516) | 6.815 (1.542) | 0.085 | 6.012 (1.424) | 6.587 (1.685) | 0.078 | 6.500 (1.009) | 7.025 (1.828) | 0.471 |
| Platelets (10<sup>9</sup>/L) | 245.738<br>(60.240) | 287.100<br>(53.450) | 0.005 | 242.351<br>(60.909) | 254.433 (58.591) | 0.354 | 276.500<br>(53.407) | 294.167 (54.621) | 0.484 |
| Red blood cells (10<sup>9</sup>/L) | 4.719 (0.654) | 5.020 (0.611) | 0.059 | 4.631 (0.679) | 4.943 (0.533) | 0.026 | 4.675 (0.534) | 5.25 0(0.565) | 0.035 |
| Lymphocytes (10<sup>9</sup>/L) | 2.026 (0.577) | 2.395 (0.510) | 0.009 | 1.949 (0.544) | 2.223 (0.622) | 0.027 | 2.188 (0.491) | 2.533 (0.494) | 0.142 |
| hemoglobin (g/L) | 141.037<br>(20.188) | 146.600 (21.01) | 0.263 | 138.558<br>(21.516) | 147.400 (14.776) | 0.041 | 143.375<br>(14.956) | 148.750 (24.647) | 0.589 |
| **Comorbidity, N (%)** |  |  |  |  |  |  |  |  |  |
| Hypertension | 12 (12) | 4 (20) | 0.280 | 8 (11) | 4 (14) | 0.736 | 0 (0) | 4 (34) | 0.094 |
| T2DM | 4 (4) | 0 (0) | >0.999 | 3 (4) | 1 (4) | >0.999 | 0 (0) | 0 (0) | >0.999 |
| MAFLD | 30 (28) | 11 (55) | 0.018 | 17 (22) | 13 (43) | 0.028 | 5 (63) | 6 (50) | 0.670 |
| **Seroconversion of neutralizing antibody to live SARS-CoV-2, N (%)** |  |  |  |  |  |  |  |  |  |
| Day 28 | 26 (25) | 2 (10) | 0.240 | 16 (21) | 10 (34) | 0.174 | 0 (0) | 2 (17) | 0.475 |
| Day 57 | 107 (100) | 20 (100) | >0.999 | 77 (100) | 30 (100) | >0.999 | 8 (100) | 12 (100) | >0.999 |
| Day 180 | 30 (29) | 11 (55) | 0.018 | 0 (0) | 30 (100) | <0.001 | 0 (0) | 12 (100) | <0.001 |
| **GMT of neutralizing antibody to live SARS-CoV-2 (AU/mL)** |  |  |  |  |  |  |  |  |  |
| Day 28 | 9.033 (5.837) | 7.41 (2.83) | 0.023 | 8.034 (4.35) | 11.596 (8.092) | 0.028 | 6.0563<br>(1.07254) | 8.3125 (3.29792) | 0.045 |
| Day 57 | 29.925 (30.425) | 27.133 (20.096) | 0.694 | 24.596 (27.2) | 43.603 (34.288) | 0.009 | 15.695<br>(6.17745) | 34.7583<br>(22.6869) | 0.034 |
| Day 180 | 8.94 (4.422) | 11.784 (4.297) | 0.009 | 6.905 (1.641) | 14.162 (5.021) | <0.001 | 8.475 (0.68498) | 13.9892<br>(4.28151) | 0.001 |

Data are shown as mean values (standard deviation in parentheses). Pearson $\chi^2$ test or Fisher's exact test were used to analyze the categorical outcomes and student t-test or Welch t-test for continuous outcomes. Hypertension was defined as systolic blood pressure $\geq 140$ or diastolic blood pressure $\geq 90$ mmHg. T2DM: Type 2 diabetes mellitus, which was defined as fasting glucose $\geq 7.0$ mmol/L. MALFD: metabolic associated fatty liver disease. Group 3: the group that became seronegative before D180; Group 4: the group that was seropositive at least until D180.

TMT-based analysis involved 528 samples, including pooled controls for aligning data from different batches to evaluate quantitative accuracy, and technical replicates for evaluating the reproducibility of the assay or technique. These samples were distributed into 33 batches from three time points: D0, D28, and D57. We quantified 7342 PBMC proteins and 1715 serum proteins (Table S2; Figures 1A and S2A). The median coefficients of variance (CV) for the pooled samples were 15.35% and 19.32% for the PBMC and the serum data, respectively (Figure S2B). The Pearson correlation coefficients of the technical replicates were 98.09% and 96.82% for PBMC and serum, respectively (Figure S2C). These results showed the robustness of our data and its relatively high consistency and reproducibility.

### Machine learning model for predicting the antibody generation

We next developed a set of models for predicting the seropositivity of individuals 57 days after their first vaccination dose and 28 days after their second one (at D57) based on the proteomics and clinical indicators collected prior to both doses (at D0). Machine learning models were developed using XGBoost (Chen and Guestrin, 2016)(Figure 2A). Proteins or clinical indicators with a significant difference (p-value < 0.05) between the two classes and with |log_2_(fold change)| > 0.25 in the discovery dataset were included in our final feature set. Then, some sparse proteins (NA rate > 50%) were also removed. We optimized the models’ parameters in the discovery dataset (Cohort 1), and generated a model based on the five PBMC proteins and another based on the seven serum proteins. Using the test cohort (Cohort 2), the PBMC model achieved an Areas Under the Curve (AUC) score of 0.84, while the serum one of 0.82 (Figure 2A). Next, we developed an ensemble model combining these two models, which led to better performance (AUC = 0.87) (Figure 2A).

**Figure 2.**
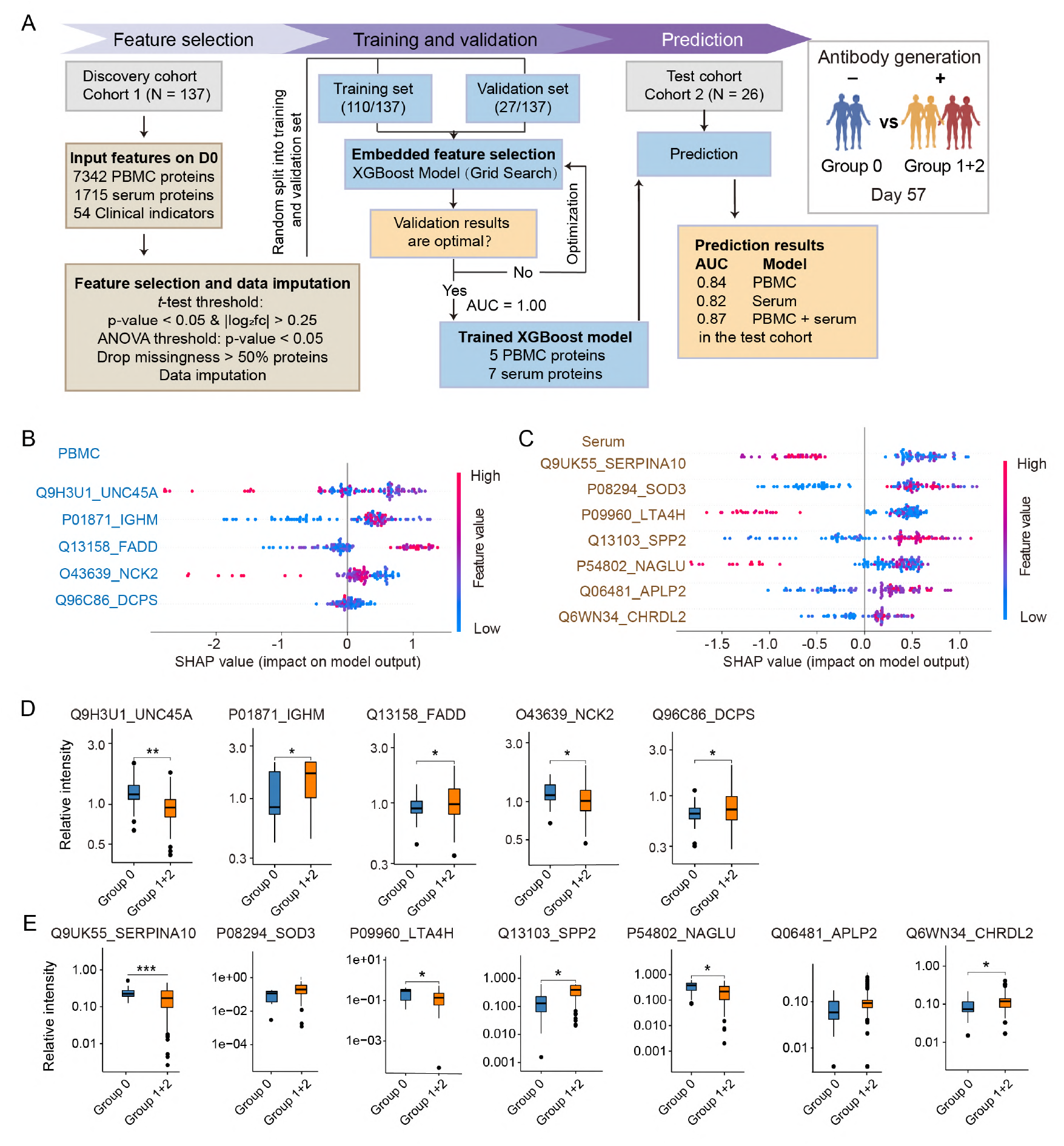
Machine learning-based prediction of individuals’ seronegative or seropositive status based on their PBMCs and serum proteins before vaccination. **(A)** Our machine learning-based predictor was based on PBMC, serum, and both types of proteins. We used the samples from a discovery cohort (Cohort 1, N = 137) to optimize the model’s parameters, the discovery dataset was randomly split into a training (80%) and a validation (20%) dataset. The model was then tested using a test cohort (Cohort 2, N = 26): the first based on PBMC biomarkers and the second on serum biomarkers. We next developed a third model that was an ensemble of the two previous ones. This third model led to an AUC of 0.87, which was higher than using PBMC or serum proteins individually. **(B)** The SHAP values of the five PBMC proteins were prioritized using the machine learning model. **(C)** The SHAP values of the seven serum proteins were prioritized using the machine learning model. **(D)** Boxplots of the selected biomarker proteins from the PBMC samples. **(E)** Boxplots of the selected biomarker proteins from the serum samples. Asterisks in **(D)** and **(E)** indicate the statistical significance based on the unpaired two-sided Welch’s t-test. Specifically, the p-values are: *, < 0.05; **, < 0.01; ***, < 0.001. Group 0: the seronegative group; Group 1+2: the seropositive group.

Five PBMC proteins (UNC45A, IGHM, FADD, NCK2, and DCPS) and seven serum proteins (SERPINA10, SOD3, LTA4H, SPP2, NAGLU, APLP2, and CHRDL2) were selected for our machine learning models. Most of the above PBMC biomarkers are expressed in immune cells, including B cells, macrophages, natural killer (NK) cells, and dendritic cells (HPA; Karlsson et al., 2021). They are thus associated with both innate and adaptive immunity (Figures 2B, 2D). In particular, UNC45A acts as a co-chaperone for HSP90 promoting progesterone receptor function in the cell, and is required for the NK cell cytotoxicity via lytic granule secretion’s control (Iizuka et al., 2015). IGHM is the constant region of immunoglobulin heavy chains and mediates the effector phase of humoral immunity, which eliminates the bound antigens. FADD is an adaptor molecule that interacts with various cell surface receptors, mediates cell apoptotic signals, and is essential in early T cell development (Kabra et al., 2001). The seven serum biomarkers are associated with immunity and metabolism (Figures 2C and 2E): SERPINA10 and SPP2 are secreted proteins associated with coagulation and metabolism; LTA4H is enriched in Kupffer cells, monocytes, and neutrophils; NAGLU is mainly expressed in most immune cells; SOD3 and CHRDL2 can interact with the extracellular matrix (ECM) organization (HPA; Karlsson et al., 2021).

Based on our three models, only two participants from the test cohort (Nos. 209 and 233) were mispredicted. Possibly, their predictions were affected by their drug treatments. Specifically, participant No. 209 was incorrectly predicted to be seronegative. This may be due to the long-term treatment with simvastatin and rosuvastatin against hyperlipidemia, which have been suggested to enhance the immune response (Guerra-De-Blas et al., 2019; Karmaus et al., 2019). Participant No. 233, whose atherosclerosis was treated with bisoprolol fumarate before vaccination, was predicted to be seropositive despite being seronegative at D57. Despite these two mispredictions, our results showed that PBMC and serum proteomics could well predict the individual host responses after vaccination. In addition, predicting those being negative at D28 and then converting (Group 1) or never converting (Group 0) could allow for the earlier switch to another vaccine. Similarly, we developed a set of models based on proteins at D0 and achieved an AUC score of 0.843 (PBMC model), 0.847 (serum model) and 0.853 (ensemble model combining these two models) to predict the individual host responses after vaccination (Figures S3A-C).

### Increased innate and adaptive immunity in the seropositive group

We next explored the differences between the seropositive and the seronegative groups using the PBMC data. Thirty-eight proteins were differentially expressed (DEPs) within the PBMC proteome between the two groups at three time points (Benjamini-Hochberg (B-H) adjusted p-value < 0.05, |log_2_(fold change)| > 0.25) (Table S3, Figures 3A and 3B). In particular, 33 proteins were dysregulated at D0 or D28. This result suggests that the immune system of the two groups was different at baseline (D0) and was most strongly activated during the early stage after vaccination.

**Figure 3.**
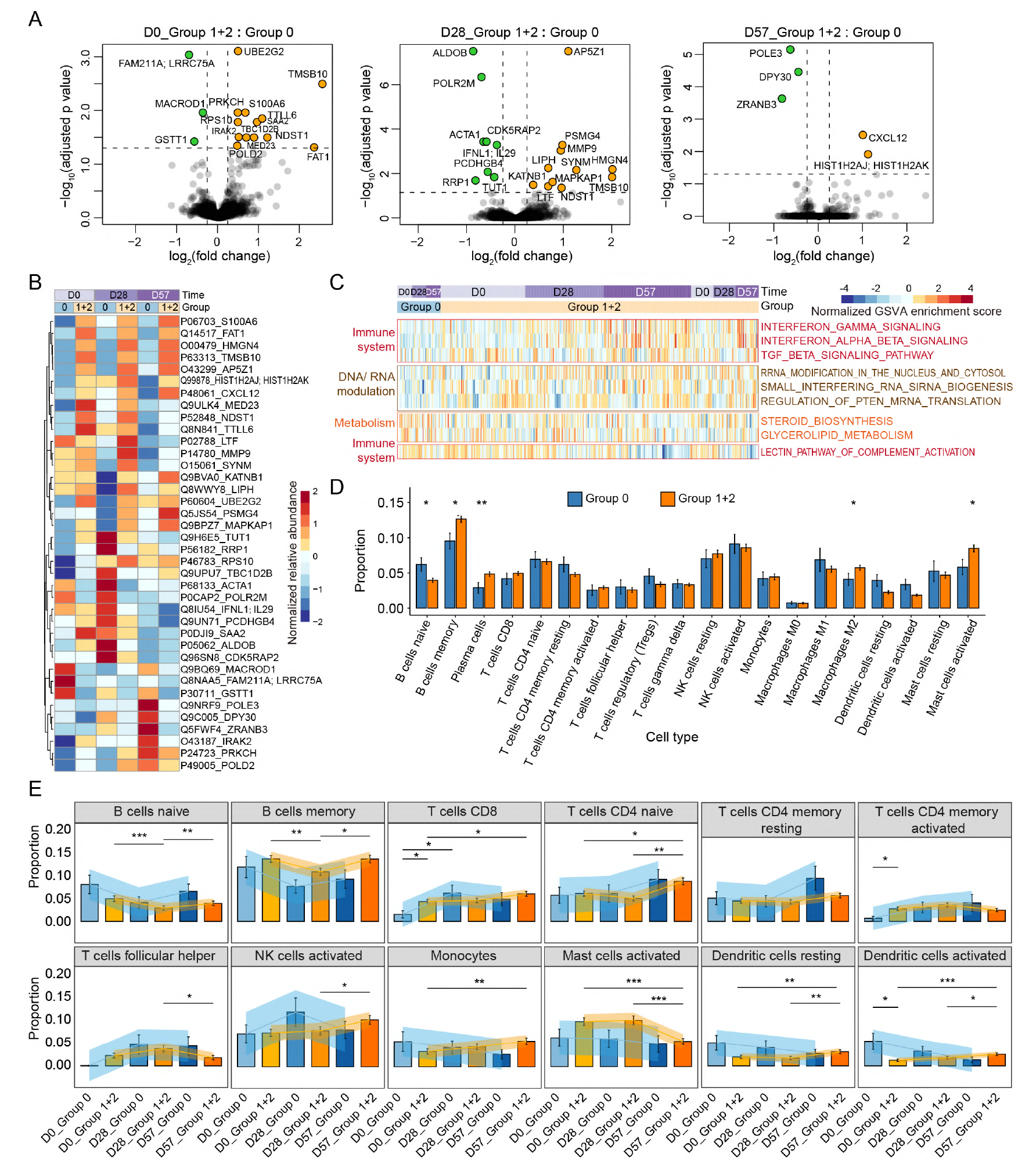
Comparison of the immune responses in the seropositive and the seronegative groups using the PBMCs proteome. **(A)** Identification of NAb status-associated proteins in PBMC using volcano plot analysis on D0, D28, and D57 (two-sided unpaired Welch’s t-test). The log_10_(B-H adjusted p-value) is plotted as a function of the log_2_(fold change) between seropositive and seronegative samples (B-H adjusted p-value < 0.05, |log_2_(fold change)| > 0.25). **(B)** Heatmap of the proteins that were significantly regulated in **(A)**. The expression of each protein is shown for both immune response groups and at D0, D28, and D57. **(C)** Heatmap of the most significantly differentially enriched pathways between the seropositive and the seronegative groups generated using GSVA (B-H adjusted p-value < 0.05, |log_2_(fold change)| > 0.25). **(D)** Barplot visualizing the inferred proportions of 20 immune cell types. The average proportions of the 20 immune cell types were derived from the seronegative (Group 0) and the seropositive (Group 1+2) groups. **(E)** Barplots visualizing the average proportions (mean ± standard error of mean) of B cells, T cells, and several innate immune cells, in the seronegative (Group 0) and seropositive (Group 1+2) groups at three time points. Asterisks in (**D**) and (**E**) indicate the statistical significance based on the Mann-Whitney rank test. P-value: *, < 0.05; **, < 0.01; ***, < 0.001.

A gene-set variation analysis (GSVA) was then used to identify the most significantly enriched pathways in the seropositive and the seronegative groups. The resulting pathways (B-H adjusted p-value < 0.05, |log_2_(fold change)| > 0.25) were mainly involved in the immune system. They included the IFNγ, IFNa and IFNb signaling, RNA and DNA modulation, and metabolic pathways. Most of these pathways were upregulated in the seropositive group (Figure 3C).

The DEPs among the three immune response groups – Group 0, Group 1, and Group 2 (Figure 1A) – were primarily involved in RNA metabolism, cellular processes, and cytoskeleton regulation-related pathways (Figures S4A-D). Therefore, the dysregulation of these proteins may have contributed to elevated immunity. This agrees with the functional analysis between the seropositive and the seronegative groups. Over 80% of DEPs observed between Group 1 and Group 0 were also detected in the comparison between seropositive (Group 1+2) and seronegative (Group 0) groups (Figures S3D, S3E and Table S3).

We next analyzed the immune cells composition of the PBMCs in our experiment using the deconvolution algorithm of CIBERSORT (Newman et al., 2015). The seropositive group showed an increase in memory B cells and a reduction of naïve B cells (Figure 3D). In addition, CD8^+^ T cells and activated memory CD4^+^ T cells were significantly higher in the seropositive group than in the seronegative one at D0. The adaptive immune responses were thus significantly enhanced at the baseline in the seropositive group (Figure 3E). At D57, the memory B cells and the CD8^+^ T cells showed an upward trend over time and were significantly higher in the seropositive group. This result is consistent with the reported host responses to SARS-CoV-2 infection and vaccination (Chen et al., 2021; Sette and Crotty, 2021). Furthermore, some innate immune cells, such as monocytes, activated NK cells, and activated dendritic cells, also increased over time in the seropositive group (Figure 3E). Moreover, the early seropositive group showed increased memory B cells, activated NK cells, M1 and M2 macrophages (Figures S4F-G). These results show that the proportion of SARS-CoV-2-specific memory lymphocytes may increase after vaccination with CoronaVac^®^.

### The interaction between metabolism and immunity is linked with seroconversion

We next investigated the differences between the seropositive and the seronegative groups using the serum data. A total of 13 DEPs were found at D0 and D28, and two at D57 (B-H adjusted p-value < 0.05, |log_2_(fold change)| > 0.25) (Table S3, Figures 4A-B). In line with our findings from the PBMC data, more DEPs were identified at D0 and D28 than at D57. All the DEPs were upregulated in the seropositive group except TTR. Misfolding and aggregation of TTR, causing amyloid thyroxine protein amyloidosis, has been reported associated with a higher risk of COVID-19 morbidity and mortality (Brannagan et al., 2021). This finding is consistent with our results, as TTR was downregulated in the seropositive group. Many of our serum DEPs are secreted proteins and include components of the immunoglobulin family: IGKV1-8, IGKV1-16, and IGHV3-15 (Schroeder and Cavacini, 2010).

**Figure 4.**
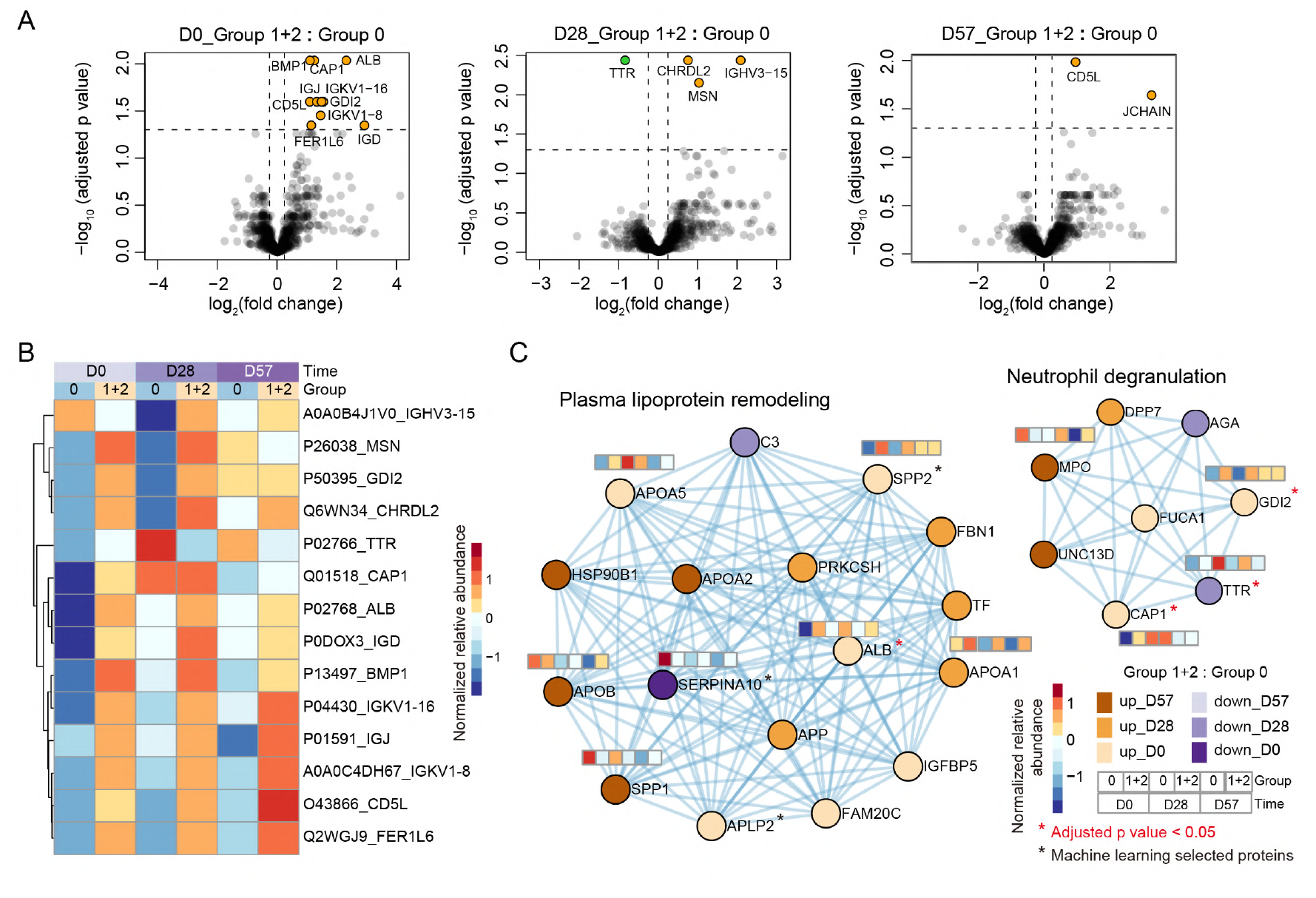
Comparison of the immune responses in the seropositive and the seronegative groups using the serum proteome. **(A)** Identification of NAb status-associated proteins in serum using volcano plot analysis on D0, D28, and D57 by the two-sided unpaired Welch’s t-test. The B-H adjusted p-values are plotted as functions of the log_2_(fold change) of the mean values between the seropositive and the seronegative samples (B-H adjusted p-value < 0.05, |log_2_(fold change)| > 0.25). **(B)** Heatmap of the proteins that were significantly regulated in **(A)**. The expression of each protein is shown for both immune response groups and at D0, D28, and D57. The most significantly enriched networks generated using significantly dysregulated proteins from the serum proteome. Proteins involved in plasma lipoprotein remodeling and neutrophil degranulation are shown with their expression levels in the seropositive and the seronegative groups at three time points. The cutoff of the dysregulated proteins was set at p-value < 0.05 and |log_2_(fold change)| > 0.25. The proteins highlighted with a red * had B-H adjusted p-values < 0.05, while those with a black * were selected from our optimized machine learning models. Group 0: the seronegative group; Group 1+2: the seropositive group.

Further functional analyses were performed on the DEPs between the seropositive and the seronegative groups, and among the three immune response groups. The significantly enriched functions were neutrophil degranulation, acute phase response signaling, and hemostasis (Figures 4B-C, S5A-B, Table S4). It has been shown that the enriched apolipoprotein family could induce the activation of leukocytes, especially the degranulation of neutrophils (Botham and Wheeler-Jones, 2013). Our analyses showed that 15 out of the 17 proteins involved in plasma lipoprotein remodeling and six out of the eight proteins involved in neutrophil degranulation were upregulated in the seropositive groups (Figure 4C). This finding suggests that the interaction of metabolism and immunity is closely linked with seroconversion.

### Predicting individual antibody persistence to guide booster shot planning

To predict whether the antibodies produced after the CoronaVac^®^ vaccination could last for at least 180 days, we generated machine learning models based on the proteomics data and the clinical indicators collected prior to vaccination. In this analysis, we excluded participants from Group 0 (the seronegative ones) and those without clinical indicators on D180. Then the remaining two cohorts (Table 2) were the discovery cohort (Cohort 3, N = 107) and the test cohort (Cohort 4, N = 20). Similar as before, proteins with a significant difference (p-value < 0.05) between two classes and with |log_2_(fold change)| > 0.25 in the discovery dataset were included in our final feature set. Then, some proteins (NA rate > 50%) were removed. We optimized the models’ parameters in the discovery dataset. An AUC score of 0.79 was obtained using only the PBMC proteins (Figure 5A), indicating that PBMC proteomics had an excellent prediction ability of the antibody response after both 57 and 180 days.

**Figure 5.**
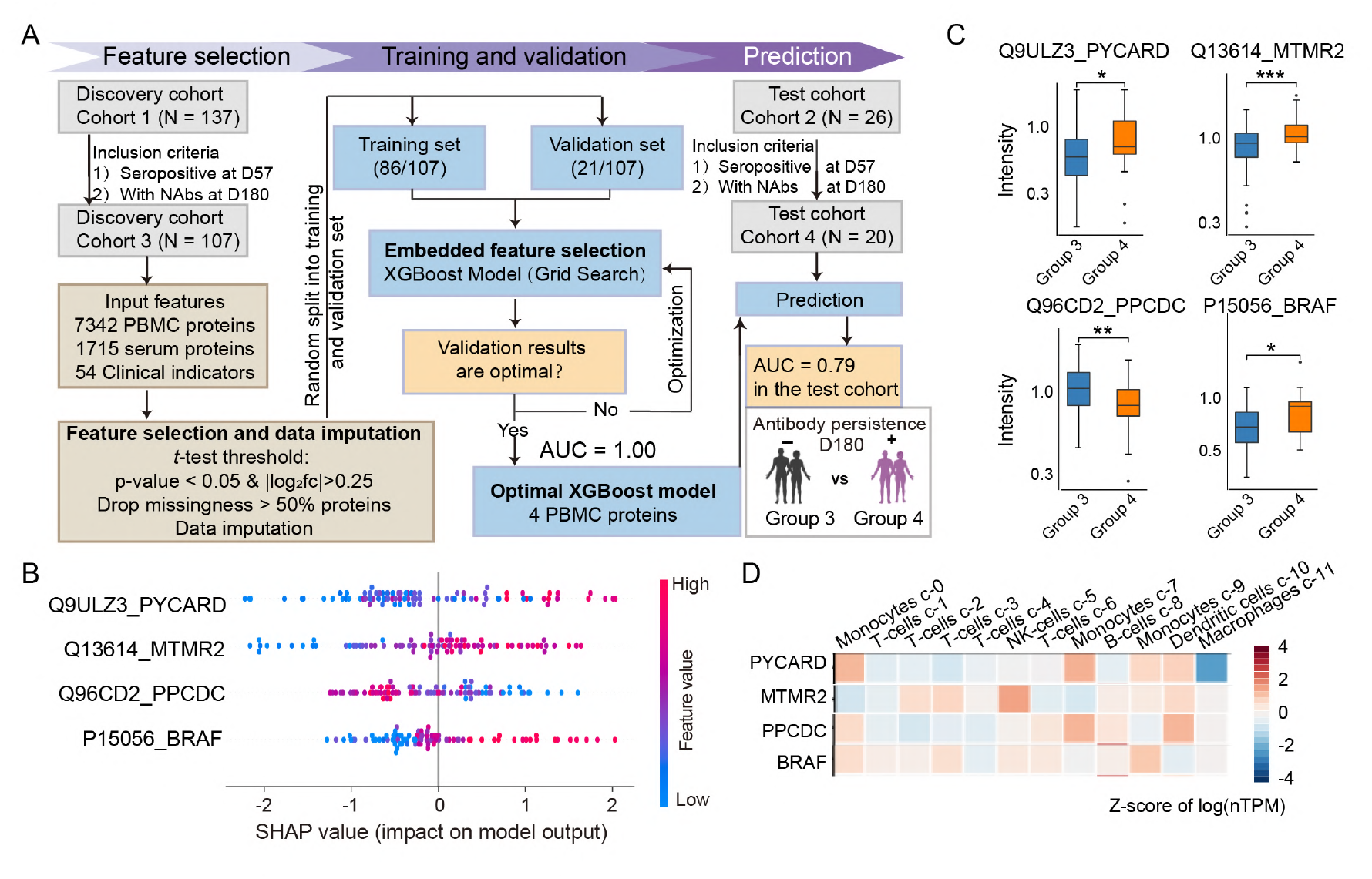
Proteomics of seropositive and seronegative individuals 180 days after CoronaVac^®^ vaccination. **(A)** Workflow for generating a model to predict the antibody persistence till D180. We discarded participants from Group 0 (the seronegative ones) and those without clinical indicators on D180, the remaining two cohorts: a training cohort (Cohort 3, N = 107) and a test cohort for the validation (Cohort 4, N = 20). **(B)** SHAP values of the machine learning classifier trained with selected PBMC proteins. **(C)** Expression of the selected proteins from the PBMC samples. The asterisks indicate the statistical significance based on the unpaired two-sided Welch’s t-test. P-value: *, < 0.05; **, < 0.01; ***, < 0.001. Group 3 (N = 107): the seronegative group on D180; Group 4 (N = 20): the persistently seropositive group. **(D)** Relative expression of the proteins selected for our model in the different cell type clusters of PBMCs (data from the Human Proteins Atlas(HPA; Karlsson et al., 2021)).

The four PBMC biomarkers (PYCARD, MTMR2, PPCDC, and BRAF) selected by machine learning showed different expression patterns of the immune cells between Groups 3 and 4 (Figure 5C). In particular, PYCARD and PPCDC are mainly expressed in innate immune cells, like monocytes and dendritic cells. MTMR2 and BRAF, on the other hand, are expressed in innate and adaptive immune cells, including NK cells, monocytes, T cells, and B cells (Figure 5D).

However, seven participants were incorrectly predicted using this model. Specifically, participants Nos. 209 and 216 were incorrectly classified, probably because they both received simvastatin and rosuvastatin (Guerra-De-Blas et al., 2019; Karmaus et al., 2019). In addition, three participants (Nos. 212, 222, and 225) with fatty liver disease and metabolic abnormalities were also wrongly predicted, probably due to their metabolic conditions. No. 226 was misclassified because of receiving dexamethasone and amoxicillin.

## Discussion

### Predicting the host response to CoronaVac^®^ vaccination using a machine learning model

We conducted a TMT-based proteomics analysis to profile the PBMC and serum features that could affect the response to the CoronaVac^®^ vaccination. Using a set of biomarkers measured before vaccination, we built three models to predict individual NAb levels at D57 and another model to predict the persistence of NAbs until at least D180. These potential biomarkers, which were used to distinguish different host responses, were validated using an independent cohort, confirming that the changes in PBMC and serum proteins reflect the pathophysiological differences between seropositive and seronegative subjects.

#### Potential biomarkers for vaccine-induced antibody generation and persistence

The proteins used by our machine learning classifiers contain several known biomarkers for COVID-19 severity or viral infections. Of note, SERPINA10, predominantly expressed in the liver and subsequently secreted into plasma, inhibits the activity of the coagulation factors Xa and XIa in the presence of protein Z, calcium, and phospholipids (Han et al., 2000). SERPINA10 is a known discriminating feature between severe and non-severe COVID-19 (Shen et al., 2020), and can be used as a classifier of disease severity (Messner et al., 2020). Specifically, it is upregulated in the severe COVID-19 cases. In our data, SERPINA10 was downregulated in the seropositive group with a negative SHapley Additive exPlanations (SHAP) value (Figures 2C and 2E). This result further highlights the role of coagulation during COVID-19 vaccination and indicates that SERPINA10 may contribute to reducing antibody generation. SOD3, an antioxidant enzyme, has been reported to be downregulated in the urine of severe COVID-19 cases (Bi et al., 2021). In our study, SOD3 was significantly upregulated in the seropositive group with a positive SHAP value, indicating that SOD3 may promote antibody generation. PYCARD, a key mediator of apoptosis and inflammation, is mainly involved in the innate immune response (Wang et al., 2017). It also contributes to T-cell immunity stimulation and cytoskeletal rearrangements coupled to chemotaxis and antigen uptake during adaptive immunity (de Souza et al., 2021). We found PYCARD was upregulated in the seropositive group with a positive SHAP value (Figures 5B-C). And thus, we suggest this protein may also promote antibody persistence. These potential biomarkers may promote or reduce antibody generation or persistence, providing therapeutic guidance for vaccination strategy.

### Mechanisms behind vaccine-induced immunity

To investigate the molecular mechanisms behind vaccine-induced immunity, we integrated our proteomics analyses and thus generated a summary of the dysregulated pathways between the seropositive and the seronegative groups (Figures 6, S6-7).

**Figure 6.**
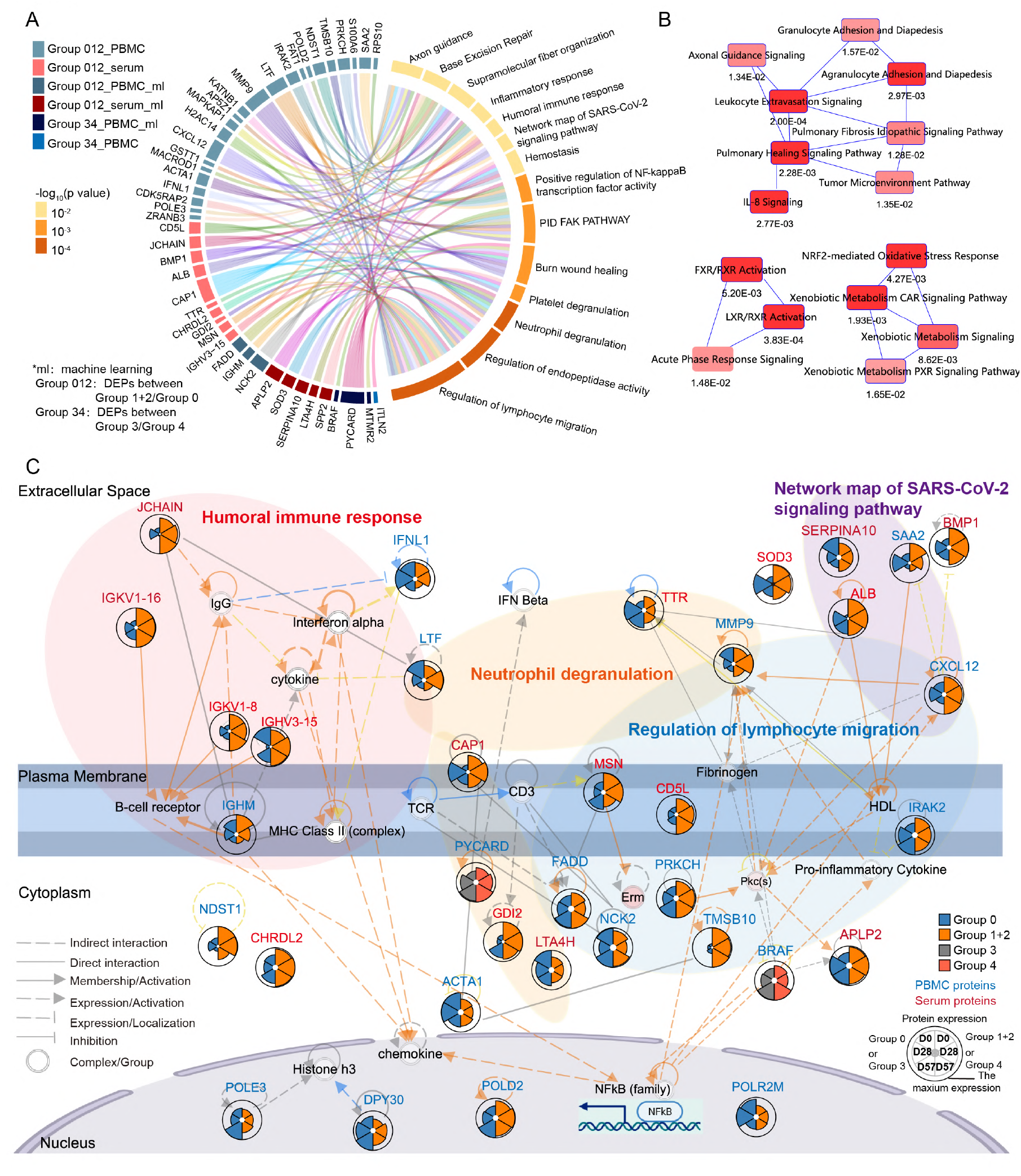
Functional and network analyses of the seropositive and seronegative groups’ immune responses: a comparison between PBMC and serum data. **(A)** Chord diagrams of the most enriched pathways based on the significantly dysregulated proteins and the potential biomarker proteins. **(B)** Network analysis of the most significantly enriched pathways (with their p-values) based on the DEPs and the potential biomarker proteins. **(C)** Key PBMC and serum proteins characterized in seronegative and seropositive recipients. Proteins involved in the humoral immune response, neutrophil degranulation, network maps of SARS-CoV-2 signaling pathways, and regulation of lymphocyte migration are shown in this network with their corresponding expression levels in the seronegative and the seropositive groups. Group 0: the seronegative group; Group 1+2: the seropositive group; Group 3: the group that became seronegative before D180; Group 4: the group that was seropositive at least until D180.

#### Neutrophil degranulation

In our data, we found several proteins involved in activating the neutrophil degranulation-based innate immunity, in particular, LTA4H, LTF, MMP9, TTR, CAP1, PYCARD, and GDI2. Specifically, MMP9, LTF, and CAP1 were upregulated at D28 in the seropositive group. Previous studies have shown that the release of MMP9 from neutrophils stimulates the migration of inflammatory cells and promotes inflammation and the degradation of the alveolar-capillary barrier (Davey et al., 2011). In our seropositive data, MMP9 was upregulated at D28 and then downregulated at D57 (Figure 6C). This result suggests MMP9 may contribute to a reduced antibody generation, and is consistent with this protein being an indicator of respiratory failure (Ueland et al., 2020) and enhanced mortality risk in COVID-19 patients (C et al., 2021). Indeed, evidence has shown that neutrophil activation is a hallmark of severe SARS-CoV-2 infection (Meizlish et al., 2021). Therefore, we speculate that a modest upregulation of neutrophil degranulation may contribute to immunity activation and *vice versa*.

#### Regulation of lymphocyte migration

Most regulators of leukocyte extravasation and lymphocyte migration were elevated at D0 or during the early stages in the seropositive group: MSN, MMP9, CXCL12, FADD, NCK2, and TMSB10. In particular, MSN interacts with members of the ezrin-radixin-moesin family and regulates lymphocyte egress from lymphoid organs (Serrador et al., 1998). TMSB10, CXCL12, and NCK2 regulate the cytoskeleton organization and are involved in transmigration (Figure 6A). By secreting proteases like MMP9, leukocytes degrade the basement membrane and penetrate the tissue interstitial spaces (Sternlicht and Werb, 2001). T-cell receptors are activated through the binding by FADD and the interaction with NCK2, consistently with our immune cell analysis (Figure 6C). Except for antigen recognition, T cell migration was positively regulated by CXCL12, FADD, and PYCARD in our seropositive groups. Intriguingly, increased FADD at baseline possibly contributed to the enhanced antibody generation at D57 (Figure 2D). Also, higher PYCARD at baseline led to a long antibody persistence based on our machine learning models (Figure 5C).

#### Humoral immune response

The B-cell receptor is a complex of surface immunoglobulin, and some of its accessory molecules, such as IGHM, IGHV3-15, IGKV1-8, and IGKV1-16, were upregulated in our seropositive group (Figure 6C). Following the receptor cross-linking, a complex cascade of signaling molecules results in NF-κB complex and B-cell receptor activation. These IgG and cytokines are expressed by JCHAIN and LTF, which were both significantly elevated in our data after vaccination. In addition, several of our overlapping proteins’ clusters of PBMC and serum were involved in neutrophil degranulation, protein-lipid complex remodeling, platelet degranulation, and complement system (Figures S6-S7). Our data show that the neutrophil degranulation-based activation of the innate immunity, the multiple immune cell migration enhancement, and the humoral immune response activation are dysregulated at baseline and during the early stages after vaccination (Figures 6A-C).

### Comparisons with other studies before/after vaccination

Several studies of COVID-19 vaccination have identified modulation of multiple proteins, metabolites, and gene expression after vaccination (Arunachalam et al., 2021; Liu et al., 2021; Zhang et al., 2021; Wang et al., 2022). However, no study has systematically investigated the heterogeneous hematological host responses to vaccination in both PBMCs and sera. Neither has any study presented any means to predict the host responses of vaccination. Vaccine-induced protection against COVID-19 may involve NAbs, T-cells, and innate immune mechanisms. In the comparative analysis of multiple vaccines, T cell responses in CoronaVac^®^ remains unclear (Sadarangani et al., 2021). Our PBMCs analysis showed that CD8^+^ T cells, memory B cells, and activated NK cells were increasingly upregulated in the seropositive group. Previous study showed that mRNA vaccinations can significantly enhance the innate immune response, as proven by the greater frequency CD14^+^ CD16^+^ inflammatory monocytes and higher concentration of plasma IFNγ (Arunachalam et al., 2021). In our PBMC proteome, monocytes and the IFNγ, IFNa and IFNb signaling were elevated in the seropositive groups. A single-cell RNA-sequencing study of the PBMCs of healthy subjects revealed that, after CoronaVac^®^ vaccination, the levels of B cells, T cells, NK cells, and myeloid cells better resembled those of COVID-19 recovery controls rather than their own before vaccination (Zhang et al., 2021). Similarly, our PBMCs analysis showed that CD8^+^ T cells, memory B cells, and activated NK cells were increasingly upregulated in the seropositive group. Consistent with other reports (Wang et al., 2022), our findings support that the humoral immune response, complement activation were induced by CoronaVac^®^. What’s unique in the seropositive participants is activated regulation of lymphocyte migration pathway, which suggests enhanced immunity.

### Planning booster shot and their benefits

Due to the relatively high effectiveness of booster immunization against severe COVID-19, hospitalization, and even the Omicron variant (Xue et al., 2022), it should be strongly supported and administered at the appropriate time. The effectiveness and the safety of boosters have been assessed via large-scale randomized studies and individuals, proving the booster’s benefits and the negligible impact of its immune-mediated side effects (Zeng et al., 2021). Boosting is particularly important for specific subpopulations: individuals who generate less or shorter-lived NAbs and those who are immunocompromised, such as our seronegative participants (Group 0). Moreover, the vaccination strategy may change for recipients with heterologous or homologous vaccinations (Costa Clemens et al., 2022). Our machine learning models predict the seropositivity of individuals at D57 and their NAbs persistence until at least D180 using potential blood-derived protein biomarkers. These tools can establish which populations or individuals may generate enough and persistent NAbs, and therefore help plan precise booster administrations. Furthermore, a better balance between primary vaccination and booster may benefit more countries in the global fight against COVID-19 (Krause et al., 2021).

In summary, we performed a systematic PBMC and serum proteomic study of the heterogeneous hematological host responses to vaccination. We developed a machine learning model based on a panel of proteins expressed at baseline to predict antibody generation and decline after vaccination. The model can be potentially used to identify the individuals of high risk, and guide booster shot, or recommendation of other vaccines. Furthermore, our data also provides a panoramic view of the molecular changes in PBMCs and serum after vaccination.

### Limitations of the study

The findings of this study have to be considered in light of some limitations. First, the predictive models need to be further validated in larger cohorts and multicenter samples, both biologically and clinically. Second, the explanations for misclassifications are not very strong, may because of complex drug history. Third, the B cell and T cell responses and the neutralization tests were analyzed in the mixed PBMCs but not assessed *in vitro*.

## Data Availability

All data are available in the manuscript or the supplementary material. The proteomics data are deposited in ProteomeXchange Consortium (https://www.iprox.org/). Project ID: IPX0004305000. All the data will be publicly released upon publication.

## Abbreviations

AGC: (automatic gain control)
AUC: (area under curve)
B-H: (Benjamini-Hochberg)
COVID-19: (coronavirus disease 2019)
CV: (coefficients of variance)
DEPs: (differentially expressed)
ECM: (extracellular matrix)
FDR: (false discovery rate)
GMT: (Geometric Mean Titers)
GSVA: (gene-set variation analysis)
IFN: (interferon)
MALFD: (metabolic associated fatty liver disease)
PBMCs: (peripheral blood mononuclear cells)
SARS-Cov-2: (severe acute respiratory syndrome coronavirus 2)
SHAP: (SHapley Additive exPlanations)
T2DM: (Type 2 diabetes mellitus)

## Acknowledgements

This work was supported by the National Natural Science Foundation of China (32200763), Key medical disciplines of Hangzhou, the Medical Health Science and Technology Project of Hangzhou municipal Health Commission (A20210205), the National Key R&D Program of China (2020YFE0202200). The Affiliated Drum Tower Hospital, Medical School of Nanjing University (2022-LCYJ-MS-08). We thank Westlake University Supercomputer Center for assistance in data generation and storage, and the Mass Spectrometry & Metabolomics Core Facility at the Center for Biomedical Research Core Facilities of Westlake University for sample analysis. We gratefully acknowledge Yi Yu for data analysis, Liqin Qian for editing the manuscript.

## Data Availability

All mass spectrometry data in this paper are available in the platform iProX at https://www.iprox.org/. Project ID: IPX0004305000. All the data will be publicly released upon publication. (URL: https://www.iprox.cn/page/PSV023.html;?url=1666845005314ZrCF;Password:ZACa)

## Author contributions

J.S., T.G., and J.L designed and supervised the project. Q.Z., Y.S., L.S. collected the samples and clinical data. X.Yi., Y.W., and X.Ye. conducted proteomics analysis. Y.W., R.S., Q.Z., and X.Yi. performed the experimental design and data interpretation. L.H., Y.W., and W.G. analyzed the proteomics data. Y.H. and Y.L. performed machine learning. Q.Z. analyzed the clinical data. Y.W., Q.Z., R.S., Y.H., H.G., J.L., T.G., and J.S. wrote the manuscript with inputs from co-authors.

## Experimental Procedures

### Experimental Design and Statistical Rationale

The overall goal was systematic investigation of host responses to COVID-19 vaccines, including the heterogeneity among the recipients, and machine learning models to predict the effectiveness of vaccination using potential biomarkers at baseline. Subject information for vaccinated recipients is summarized in Tables 1-2, Table S1. Study design of the TMT-Pro labeling-based quantitative proteomics analysis of the PBMCs and sera samples is depicted in Figure 1A and S2A.

#### Participants and Samples

We recruited 163 vaccination recipients (>18 years) who were not infected with SARS-CoV-2 and some of them had stable chronic medical conditions, including hypertension, T2DM, and metabolic fatty liver disease, were eligible to be enrolled from the affiliated hospital of Hangzhou Normal University between January and February 2021, including a discovery (N = 137) and an independent test cohort (N = 26). All participants received two doses of CoronaVac^®^ (0.5 mL/dose, Sinovac life science, Beijing, China), an inactivated vaccine against SARS-CoV-2; the second dose 28 days after the first one. Blood samples were collected before vaccination (D0), then 28 (D28), 57 (D57). Blood mononuclear cells and serum were extracted from the blood samples. The xenoreactivity was also measured at D0, D28, D57 and 180 days after the first dose vaccination (D180). The NAbs for the receptor-binding domain of the SARS-CoV-2 spike protein were detected using the iFlash 2019-nCoV NAb assay (SHENZHEN YHLO BIOTECH CO., LTD, Shenzhen, China, Cat#C86109), which is a paramagnetic particle chemiluminescent immunoassay for the qualitative detection of SARS-CoV-2 NAbs in human serum and plasma using the automated iFlash immunoassay system; the cut-off value for the antibody was 10.00 AU/mL.

The participants were classified into three groups based on the xenoreactivity of their NAbs on D28 and Day 57. Specifically, Group 0 included the participants that were seronegative on D28 and D57; Group 1 included the participants that were seronegative on D28 but were seropositive on D57; Group 2 included the participants that were seropositive on D28 and D57. Groups 1 and 2 were then merged into Group 1+2 (all the seropositive participants). Group 1+2 was then split into Group 3 (seronegative at D180) and Group 4 (seropositive at D180).

This research was approved by the ethical committee of the Affiliated Hospital of Hangzhou Normal University and Westlake University (Hangzhou, China). The study was registered in the Chinese Clinical Trial Register (ChiCTR2100042717), and all participants signed a written informed consent before enrolment.

### Serum and PBMC Protein Extraction and Digestion

From each sample, 4 μL of serum were depleted of 14 high abundant serum proteins using a human affinity depletion resin (Thermo Fisher Scientific™, San Jose, USA) and then concentrated into 50 μL through a 3K MWCO filtering unit (Thermo Fisher Scientific™, San Jose, USA). More details can be found in the manufacturer’s protocols. The resulting serum samples were then prepared for mass spectrometry as described (Shen et al., 2020). Briefly, they were denatured in 8 M urea at 31.5°C for 30 min. Next, the proteins were reduced with 10 mM tris (2-carboxyethyl) phosphine (TCEP) and then alkylated with 40 mM iodoacetamide (IAA). Finally, the protein extracts were diluted and digested using a double step trypsinization for 16 hours totally (Hualishi Tech. Ltd, Beijing, China).

PBMCs were prepared as previously described (Gao et al., 2020). Briefly, 30 μL of lysis buffer in 100 mM TEAB with 20 mM TCEP, and 40 mM IAA were added to the PCT-Microtubes for 60 min. The proteins were digested using a mixture of trypsin and Lys-C for 100 min. Then, the digestion was arrested by adding 10% trifluoroacetic acid (TFA).

### LC-MS/MS Analysis

The proteome analysis was performed similar as previously described (Shen et al., 2020). Digested peptides were cleaned-up and labeled using TMTpro 16plex label reagents (Thermo Fisher Scientific, San Jose, USA). Peptides were separated into 30 fractions, which were later combined into 15 fractions. Subsequently, the fractions were dried, redissolved in 2% ACN/0.1% formic acid. All the samples were analyzed using liquid chromatography (LC)-coupled tandem mass spectrometry (MS/MS) with a data-dependent acquisition mode on an Orbitrap 480 (Thermo Fisher Scientific, San Jose, USA). During each acquisition, peptides were analyzed using a 30 minutes-long LC gradient (from 7% to 30% buffer B). The m/z range of MS1 was 375-1800, with a resolution of 60,000, normalized Automatic Gain Control (AGC) target of 300%, maximum ion injection time (max IT) of 50 ms, and compensation voltages of -48V and -68V for FAIMS Pro™. MS/MS experiments were performed with a resolution of 30,000, normalized AGC target of 200%, and 86 ms max IT for Serum and 100 ms for PBMC. The turbo-TMT and the advanced peak determination were enabled.

### Database Search for Proteomics Quantification

The mass spectrometric data were analyzed using Proteome Discoverer (Version 2.4.0.305, Thermo Fisher Scientific) and the *Homo sapiens* protein database downloaded from UniProtKB on 27 April 2020 (Fasta file containing 20,301 reviewed protein sequences). The database search was performed as previously described (Shen et al., 2020), including Carbamidomethyl (C) as a fixed modification and oxidation (M) as a variable modification. The false discovery rate (FDR) was set as 0.01. Data normalization was performed against the total peptide amount. Other parameters followed the default setup.

### Quality Control of the Proteome Data

The quality of the proteomics data was ensured at multiple levels. A pool of samples labeled by TMTpro-134N was used as the control for aligning the data from different batches. Also, we assessed the reproducibility of the data using technical replicates, water samples (buffer A) as blanks every four injections to avoid carry-over.

After removing the proteins with over 90% missing values, 6331 proteins of PBMC and 961 of serum underwent quality controls. We then assessed the coefficient of variation in the pooled samples (Figure S2B). Finally, the Pearson’s correlation values of the technical replicates (17 PBMC samples and three serum samples) were used to evaluate the reproducibility of the data (Figure S2C).

### Statistical Analysis of Clinical Indicators

Continuous variables were calculated by student t-test or Welch t-test, Pearson χ^2^ test or Fisher’s exact test for the analysis of categorical outcomes. We calculated Geometric Mean Titers (GMT) of the neutralizing antibody titers and the overall anti-Spike IgG levels, using the t-test method to compare the difference. Statistical analysis was performed by IBM SPSS Statistics 26 (Armonk, NY: IBM Corp).

### Differential Expression Analysis

A set of statistical tools were used to process and analyze our proteomics data. First, the batch effect of the serum proteome was removed using the R package combat (https://lifeinfor.shinyapps.io/batchserver/). No other significant batch effect was highlighted by principal component analysis (Figures S2D-E). For comparing the protein expressions between groups, the log_2_(fold change) was calculated using the mean values of each group. A two-sided unpaired Welch’s t-test was performed for each group pair. A one-way analysis of variance (ANOVA) was performed among three groups at three time points. Finally, the adjusted p-values were calculated using the B-H correction.

DEPs were selected by imposing the B-H adjusted p-values to be less than 0.05 and the absolute log_2_(fold change) larger than 0.25. Next, a soft clustering of the time series data was performed using MFuzz (version 2.48.0). We clustered the PBMC and serum DEPs expression along time using default settings (Figure S6). The single-cell RNA expression of PBMCs was derived from the Human Proteins Atlas (HPA; Karlsson et al., 2021).

### Estimation of the Immune Cell Type Fractions

CIBERSORT (https://cibersort.stanford.edu/) is an analytical tool for estimating the cell composition of tissues using their gene expression profiles (Newman et al., 2015). In CIBERSORT, the relative amounts of 20 human immune cell types (including naïve and memory B cells, seven T cell types, NK cells, plasma cells, monocytes, etc.) were estimated in our PBMC bulk cells using the leukocyte gene signature matrix. In addition, vaccinated individuals were divided into seronegative and seropositive groups, and the fraction of each immune cell type was investigated and visualized with bar plots using R software (R 4.0.5).

### Machine Learning

For prediction of NAbs generation on D57, we used the samples from a discovery cohort (Cohort 1, N = 137) to optimize the model’s parameters, the discovery dataset was randomly split into a training (80%) and a validation (20%) dataset. To establish the features for our machine learning models, we used a differential protein expression analysis which returned a set of biomarkers from the PBMCs and the serum (Figure 2A and 5A). Proteins with a significant difference (p-value < 0.05) between two classes and with |log_2_(fold change)| > 0.25 in the training dataset were included in our final feature set. Then, sparse proteins (NA rate > 50%) were removed. The missing values were imputed with the minimum of each protein. We decided on the top N best features as the final feature set for our model, as well as the optimal parameters, by searching for the highest AUC in the validation dataset. All individual models for these two tasks can achieve an AUC of 1.0 in the validation dataset. Finally, the results illustrated in this paper were derived from the model with the best features and parameters. The implementation of machine learning was done using Python 3.8.10 and xgboost 1.4.2 python package (Chen and Guestrin, 2016).The model was then tested using an independent test cohort (Cohort 2, N = 26): the first based on PBMC biomarkers and the second on serum biomarkers. We next developed a third model that was an ensemble of the two previous ones. This third model led to an AUC of 0.87, which was higher than using PBMC or serum proteins individually.

For prediction of NAbs persistence till D180, we discarded participants from Group 0 (the seronegative ones) and those without clinical indicators on D180, and the remaining two cohorts: a training cohort (Cohort 3, N = 107) and a test cohort for the validation (Cohort 4, N = 20). We optimized the models’ parameters in the training and a validation dataset. Similarly, we tested in Cohort 4, and an AUC score of 0.79 was obtained using only the PBMC proteins (Figure 5A).

### Functional Analyses

Specifically, we investigated 38 PBMC DEPs and 14 serum DEPs from different immune response groups using a two-sided unpaired Welch’s t-test, and 985 DEPs from PBMCs and 129 DEPs from serum were evaluated using the ANOVA test, biomarker proteins were also included. Several pathway analysis tools were used to perform the functional analysis of our significantly DEPs. Enrichments analyses based on Gene Ontology processes, KEGG pathways, Reactome gene sets, and Wiki pathways were performed using the web-based platform of Metascape (Zhou et al., 2019). With an ingenuine pathway analysis (Kramer et al., 2014) of the regulated proteins, we identified the most significantly regulated pathways; p-values were based on a right-tailed Fisher’s Exact Test, and the enriched pathways’ overall activation/inhibition state was predicted using the z-score. A pathway’s regulation was significant if its p-value < 0.05. Gene Set Variation Analysis (GSVA) was performed using the R package GSVA (version 3.11) (Hanzelmann et al., 2013) to identify the most dysregulated pathways (Canonical pathways) between the seronegative and the seropositive groups (B-H adjusted p-value < 0.05). The functional network images generated by Metascape were visualized with Cytoscape (version 3.9.0) to generate the network of predicted associations for a specific group of proteins (Otasek et al., 2019).

## Figure legends

**Figure S1.**
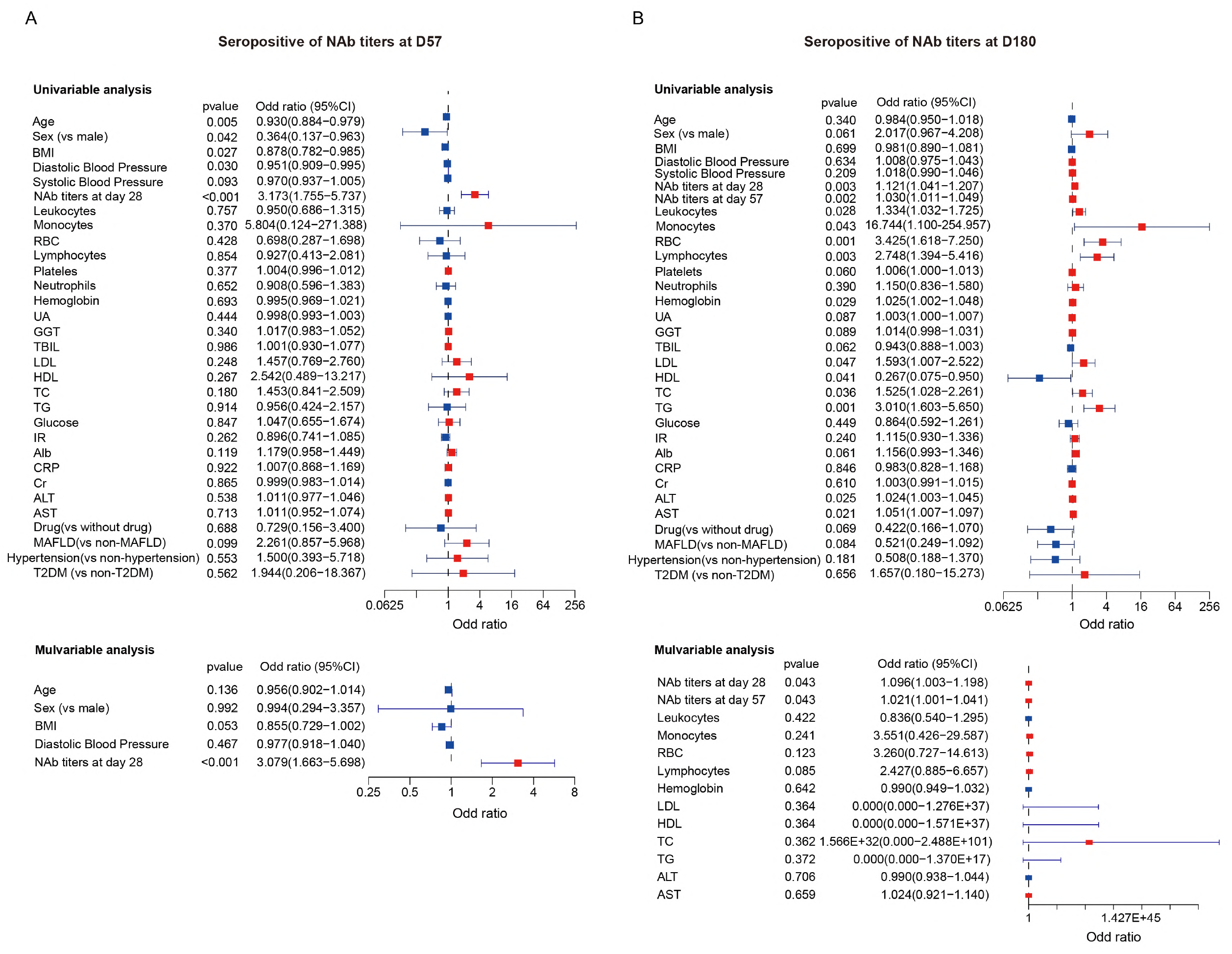
Factors associated with seropositivity of neutralizing antibodies at D57 and D180, respectively. **(A)** Multivariable logistic regression analysis was used to investigate factors associated with the generation of NAbs at D57 after adjusting sex, BMI and diastolic bold pressure. **(B)** Multivariable logistic regression analysis was used to investigate factors associated with the persistence of NAbs at D180 after adjusting leukocytes, monocytes, RBC, lymphocytes, hemoglobin, LDL, HDL, TG, ALT, and AST.

**Figure S2.**
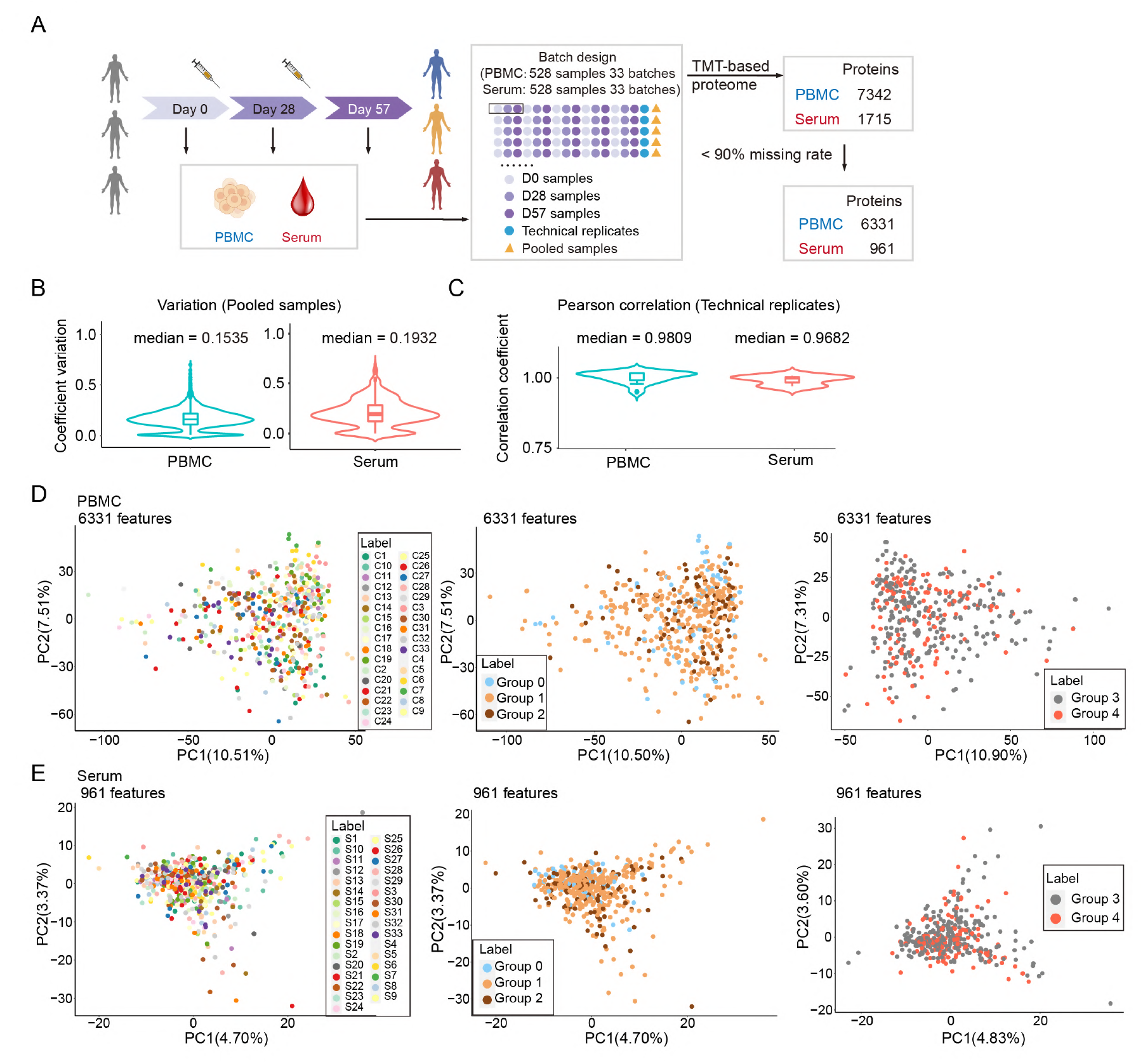
Quality controls of the proteomics data. **(A)** Study design of our TMT-labeling-based quantitative proteomics analysis of PBMCs and sera samples. Together, PBMCs and serum generated 528 peptide samples, including 33 pooled controls, distributed into 33 batches and analyzed using TMTpro 16-plex labeling based proteomics. **(B)** The proteomics data’s coefficients of variation were calculated using the abundance of the quantified proteins in the 33 pooled controls of the 33 batches. Also, they were computed after removing the outliers. **(C)** Quality control of the technical replicates based on the Pearson correlation coefficients. **(D-E)** PCA analyses were performed on all the proteomics data derived from the PBMC **(D)** and serum samples **(E)**, for three immune response groups on D57 (Group 0, Group 1, and Group 2) and two immune response groups on D180 (Group 3 and Group 4). Group 1: the late seropositive group; Group 2: the early seropositive group.

**Figure S3.**
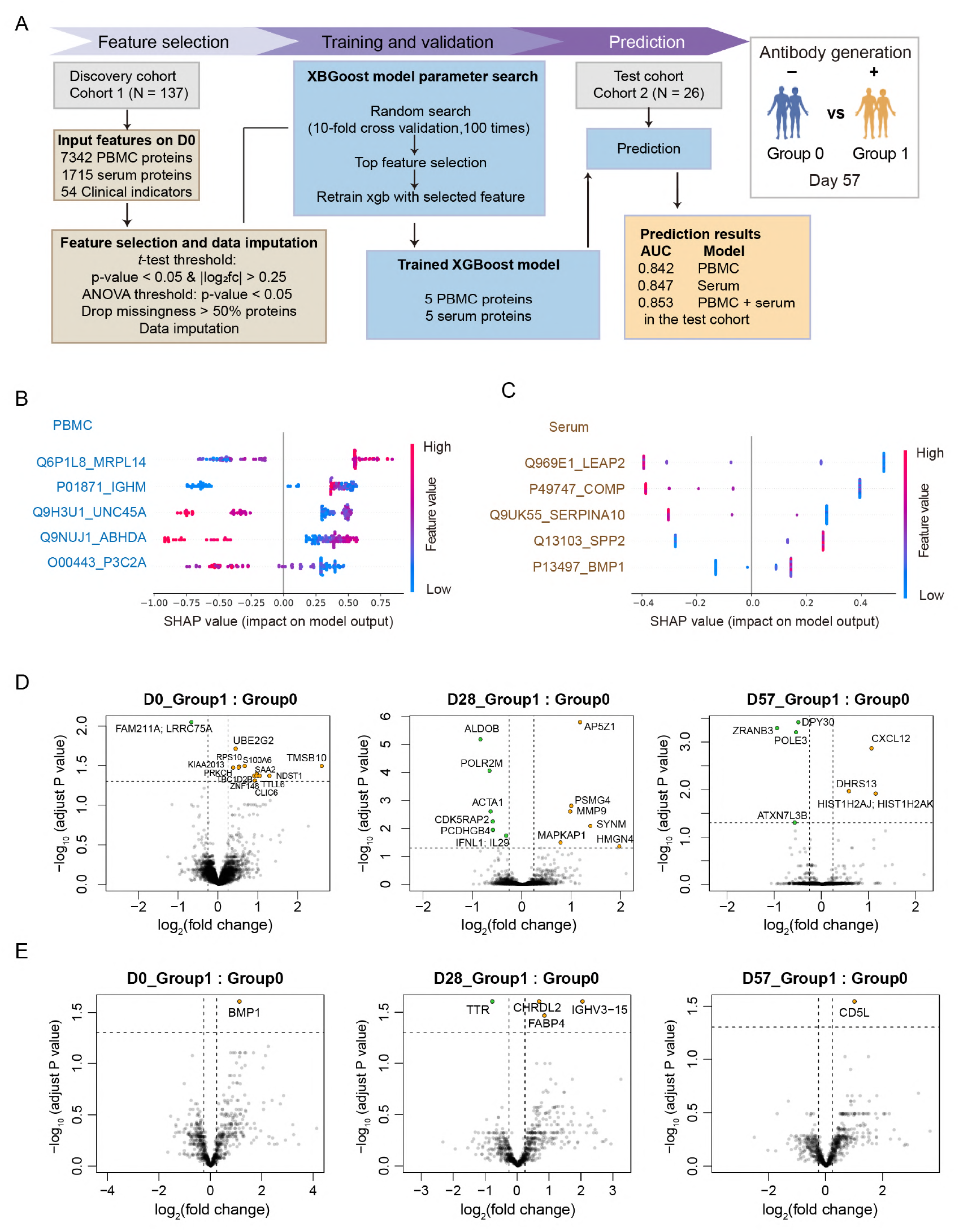
Machine learning-based prediction of Group 1 (being negative at D28 and then converting) and Group 0 (being negative at D28 and never converting) before vaccination. **(A)** Our machine learning-based predictor was based on PBMC, serum, and both types of proteins. We used the samples from a discovery cohort (Cohort 1, N = 137) to optimize the model’s parameters. The model was then tested using a test cohort (Cohort 2, N = 26): the first based on PBMC biomarkers and the second on serum biomarkers. We next developed a third model that was an ensemble of the two previous ones. This third model led to an AUC of 0.853, which was higher than using PBMC or serum proteins individually. **(B)** The SHAP values of the five PBMC proteins were prioritized using the machine learning model. **(C)** The SHAP values of the five serum proteins were prioritized using the machine learning model. Identification of NAb status-associated proteins in PBMC **(D)** and serum **(E)** using volcano plot analysis on D0, D28, and D57 (two-sided unpaired Welch’s t-test). The log10 (B-H adjusted p-value) is plotted as a function of the log2(fold change) between Group 1 and Group 0 samples (B-H adjusted p-value < 0.05, |log2(fold change)| > 0.25).

**Figure S4.**
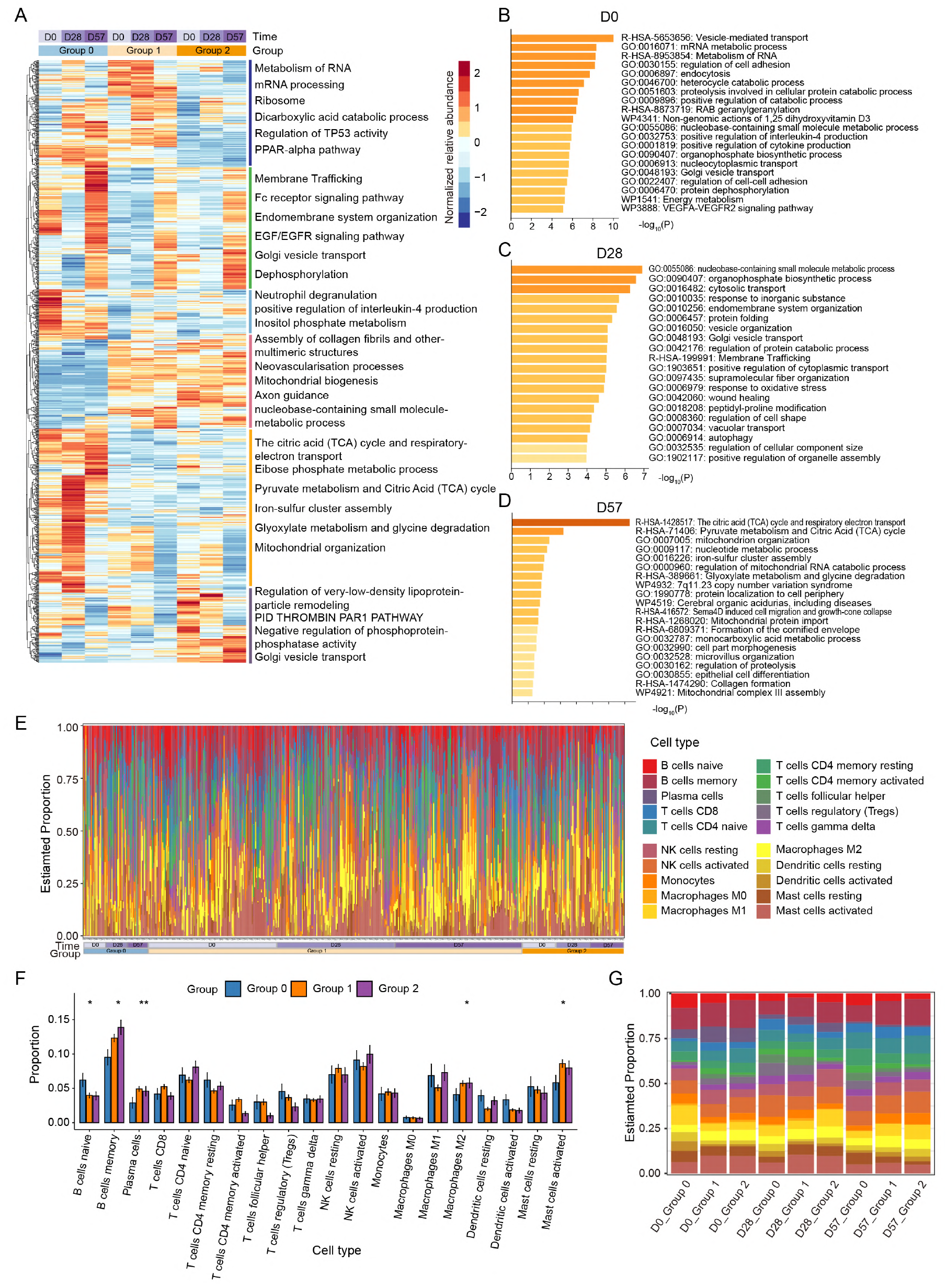
Immune response and pathway analyses using the PBMC data. **(A)** Heatmap of 985 proteins selected using the ANOVA test. Each protein is compared among the three immune response groups (Group 0, 1, and 2) on D0, D28, and D57 (p-value < 0.05). Group 0: the seronegative group; Group 1: the late seropositive group; Group 2: the early seropositive group. **(B-D)** Pathways’ enrichment based on the proteins selected using the ANOVA test to compare the three immune response groups on D0 (B), D28 (C), and D57 (D) using Metascape (log_10_ p-value). **(E)** Barplots visualizing the estimated proportions of 20 immune cell types in each PBMC sample. Each column represents a sample; the colors indicate the inferred immune cell components. **(F)** Average proportions of 20 immune cell types, in Group 0, Group 1, and Group 2, at three time points. The asterisks indicate the statistical significance based on the Kruskal-Wallis test. P-value: *, < 0.05; **, < 0.01; ***, < 0.001. **(G)** Barplots visualizing the estimated proportions of 20 immune cell types, in Group 0, Group 1, and Group 2, at three time points. Different colors indicate the predicted composition of immune cell types.

**Figure S5.**
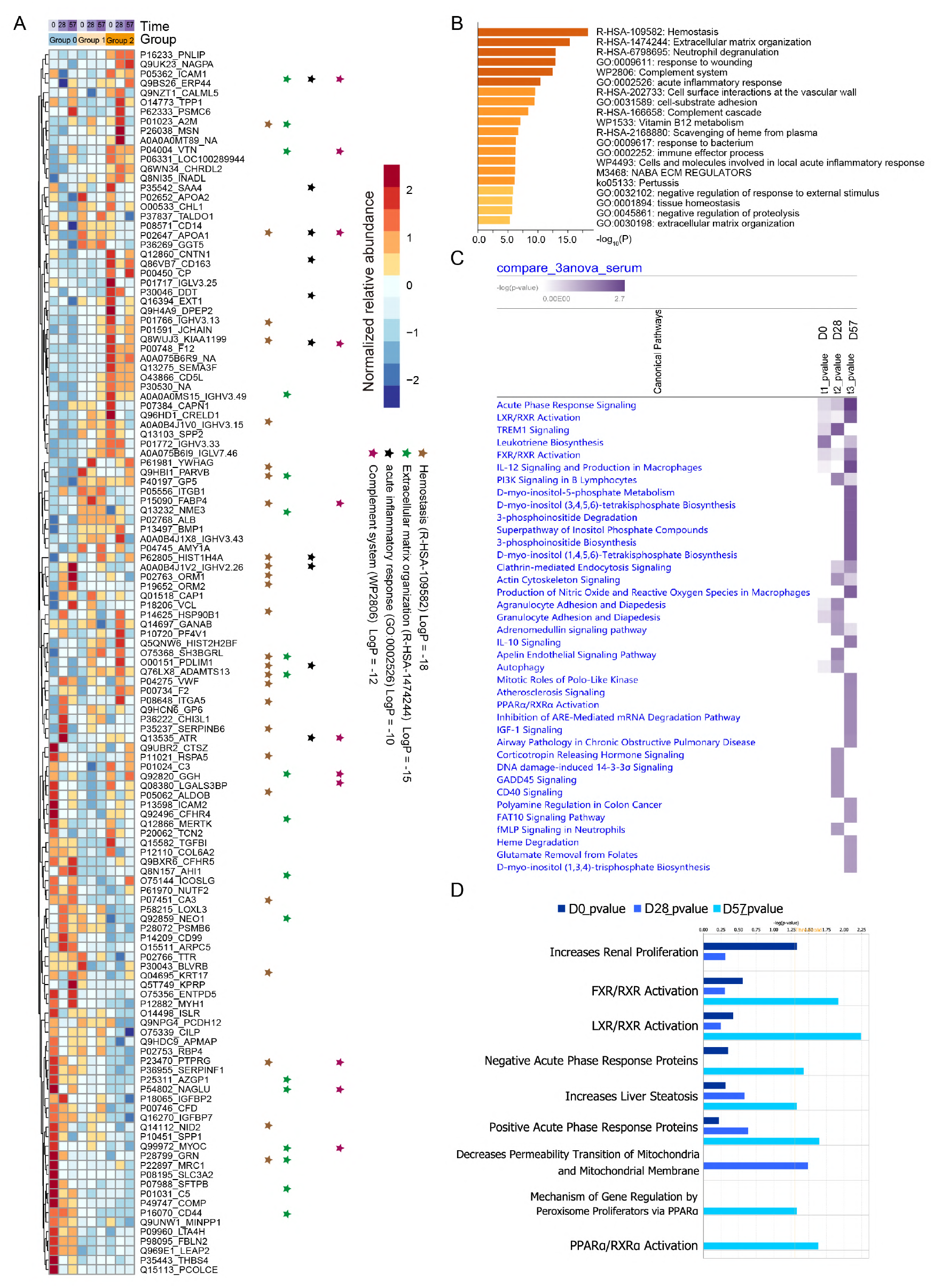
Immune response and pathway analysis using the serum data. **(A)** Heatmap of 129 proteins selected using the ANOVA test. Each protein is compared between the three immune response groups (Group 0, 1, and 2) on D0, D28, and D57 (P-value < 0.05). Group 0: the seronegative group; Group 1: the late seropositive group; Group 2: the early seropositive group. **(B)** Pathways’ enrichment based on the proteins selected in **(A)** using Metascape. **(C-D)** Comparison among three immune response groups (Group 0, 1, and 2) of the canonical pathways of proteins in (A) by IPA (Kramer et al., 2014).

**Figure S6.**
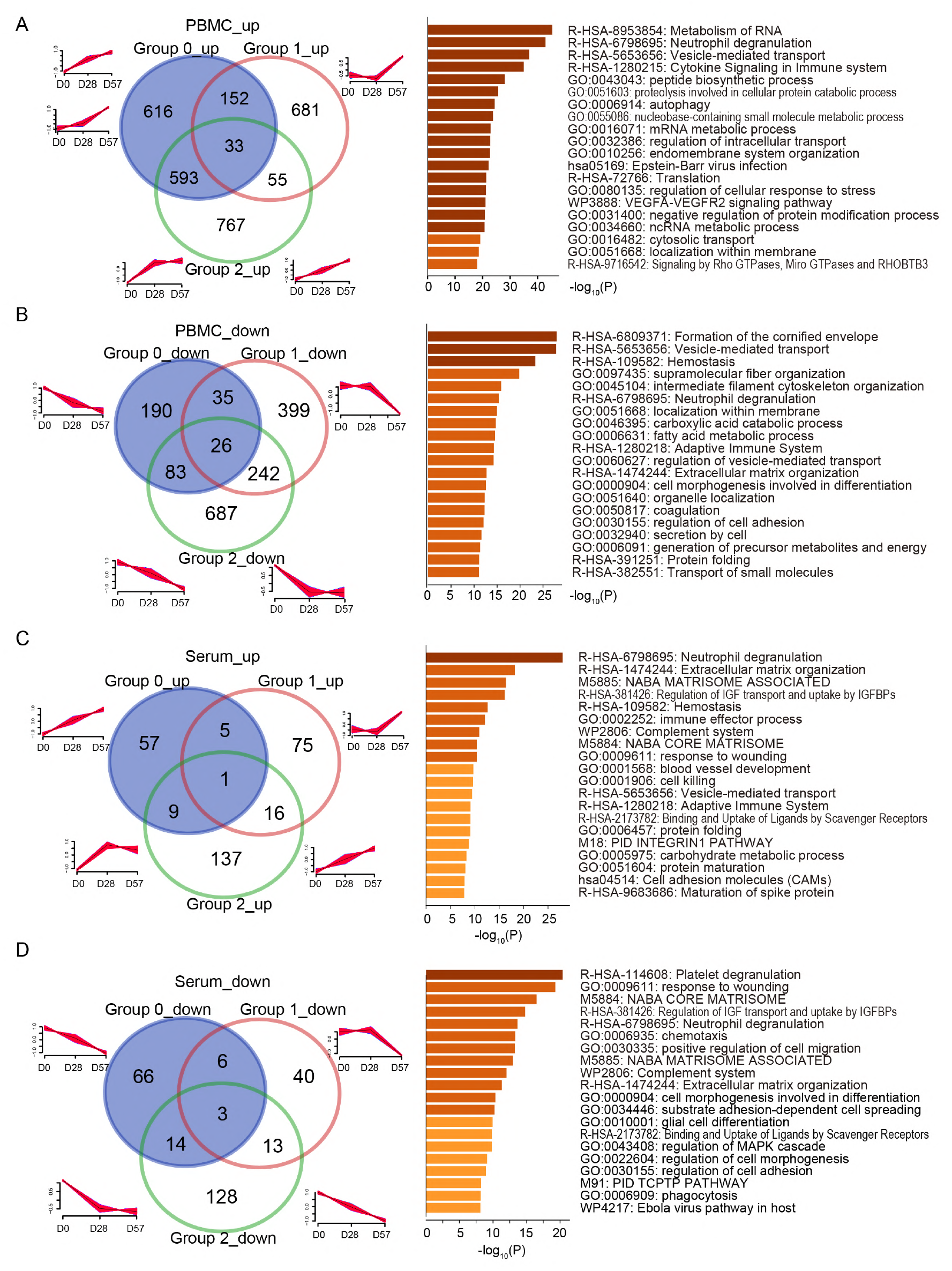
Immune response and pathway analysis using the PBMC and the serum data. **(A-B)** Pathways’ enrichment analysis of selected PBMC proteins. Specifically, 6331 PBMC proteins and 961 serum proteins were grouped into 12 discrete clusters using mFuzz, respectively. This analysis included all the proteins from the PBMC dataset that steadily increased **(A)** or decreased **(B)** over time. And the serum dataset that steadily increased **(C)** or decreased **(D)** over time. The 20 most significantly enriched pathways involving these DEPs were analyzed using Metascape.

**Figure S7.**
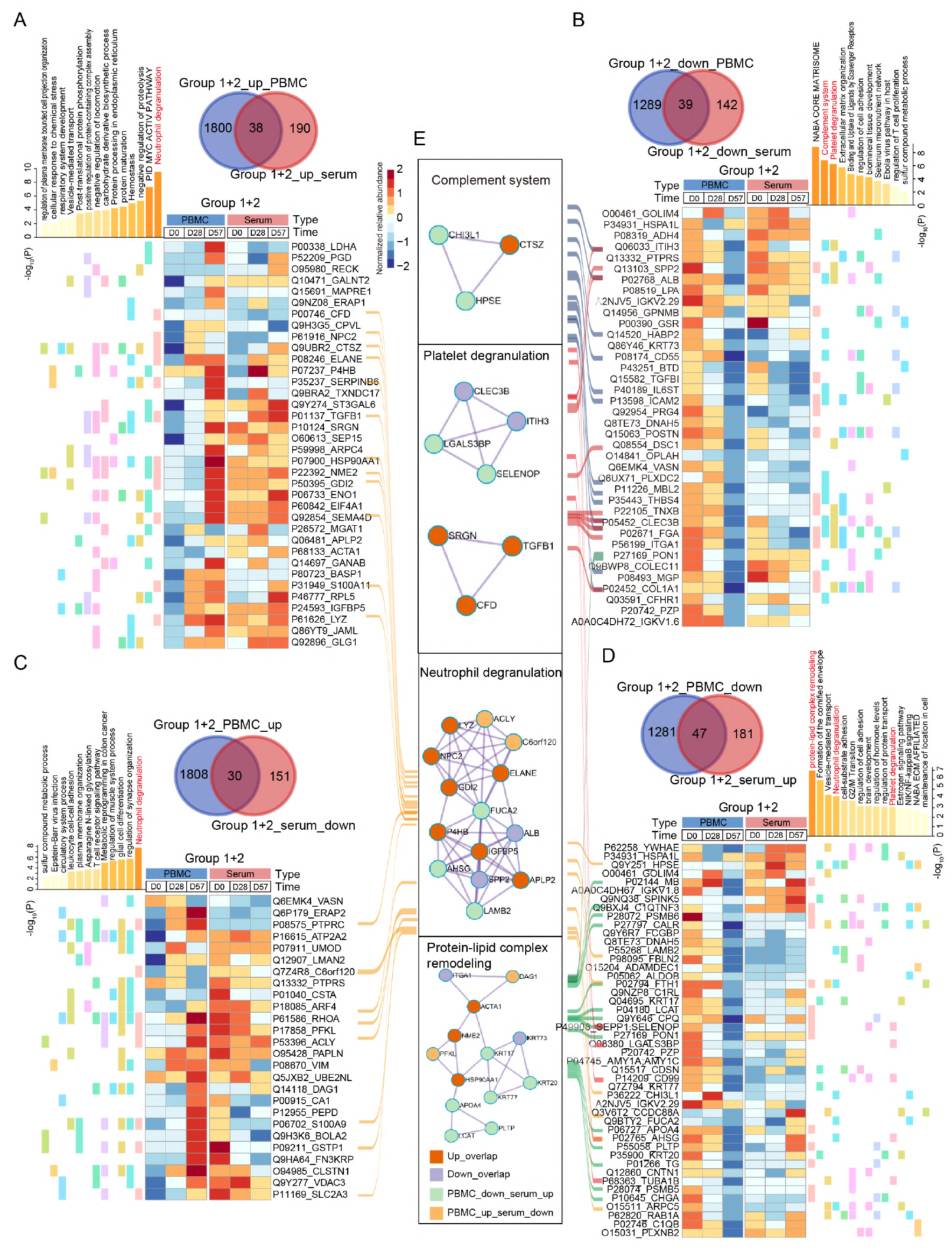
Immune response and network analysis of the seropositive groups: comparison between PBMC and serum data. The most significantly dysregulated proteins identified using mFuzz from the following groups are here compared: **(A)** upregulated in both PBMC and serum, **(B)** downregulated in both PBMC and serum, **(C)** upregulated in PBMC and downregulated in serum, and **(D)** downregulated in PBMC and upregulated in serum. The Venn diagrams show the overlaps between the PBMC (blue) and the serum proteins (red). Pathways and heatmaps were generated from the overlapping proteins from each pair. **(E)** The most significantly enriched networks generated using the DEPs from **(A-D)**. The proteins involved in the complement system, including platelet degranulation, neutrophil degranulation, and protein-lipid complex remodeling.

